# Effective connectivity of the insula as measured by cortico-cortical evoked potentials

**DOI:** 10.64898/2026.02.16.26344827

**Authors:** Cristiana Pinheiro, Maciej Jedynak, Sofia Avalos-Alais, Anthony Boyer, Julien Bastin, Philippe Kahane, Olivier David, F-TRACT Consortium

**Affiliations:** Aix Marseille Univ, INSERM, INS, Inst Neurosci Syst, 13005 Marseille, France; Univ. Grenoble Alpes, Inserm, U1216, CHU Grenoble Alpes, Grenoble Institut Neurosciences, 38000 Grenoble, France; Neurology Department, University Hospital of Grenoble, Grenoble, France; Fondation Lenval, Department of Pediatric Neurosurgery, Nice, France

**Keywords:** CCEP, directionality, effective connectivity, human brain mapping, insula, SEEG

## Abstract

Effective connectivity of the human insula, mainly assessed at rest using cortico-cortical evoked potentials (CCEPs), is not yet fully characterized at high-resolution. Here, we significantly extend prior CCEP studies of the insula by leveraging an extensive multicenter CCEP database and fine-grained anatomical atlases of the insula.

We analyzed CCEP datasets from 897 patients with refractory focal epilepsy (459 females, age: 26±14 years) explored by stereo electroencephalography and with at least one electrode contact in the insula. Efferent and afferent effective connectivity measures of nineteen insular subregions with the rest of the brain were derived at the population level, by pooling statistical properties of early brain responses to electrical stimulation pulses, as defined by the first significant component of CCEPs occurring before 100 ms. In addition, the median peak delay of the responses was measured as a proxy of the directness of the connections.

Results revealed predominant ipsilateral insular connections with frontal, parietal, central, temporal, and limbic systems. Some directional biases were observed, with more afferent connections from the caudal part of frontal lobe, central regions, temporoparietal junction, temporal pole and amygdala, and more efferent connections to the rostral part of frontal lobe, anterior cingulate, parahippocampal cortex, and hippocampus. Subregional analysis revealed a remarkably preserved topological pattern with a gradient of effective connectivity along anterior-posterior and superior-inferior axes. Along the anterior-posterior axis, the posterior insula demonstrated predominant connections with parietal, central, temporal, and limbic systems, while the anterior insula was additionally connected with the frontal system. Along the superior-inferior axis, superior insula was mainly connected with frontal, parietal, central, temporal, and limbic systems, whereas the inferior insula was primarily connected with temporal and limbic regions. Median peak delays range from 14 to 51 ms, with the fastest responses in insula surrounding areas.

This study provides the highest-resolution effective connectivity mapping of the human insula from neurophysiological data. It complements well previous structural studies with additional dynamical and causal information, and definitely establishes the insula as a topologically organised hub connecting the different brain lobes.

## Introduction

*Insula* is Latin for island, being a brain region anatomically located nestled behind the frontal, temporal, and parietal opercular cortices^1^, covering 2% of the cerebral cortex^2–6^. Architectonically, the human insula is divided into an agranular, granular, and dysgranular regions^7^. Anatomically, it is divided by the central sulcus into anterior and posterior regions. Anterior region includes anterior, middle, and posterior short gyri while posterior region comprises anterior and posterior long gyri^2^. This anterior-posterior anatomical division progressively shifts from agranular to dysgranular and then granular cortices^8^.

Current literature claim insula as being “the hub of brain”^9^ once it is implicated in a vast number of functions, namely, multisensory (viscerosensory, somatosensory, gustatory, olfactory, auditory, visual, vestibular), motor, language, cognition, memory, social emotional processing, and risky decision making^2,3,10^. As being integrated in such a wide array of functions, there is prompted interest to dissect precisely how insula influences different whole brain networks^11^. Structural connectivity analyses from diffusion MRI tractography at high-resolution^12,13^, which considered human insula subdivision in 19 regions, effectively demonstrated a clearly organised structural connectivity across insular subregions, with 1) posterior portion connected with frontal, parietal, temporal, and occipital lobes; 2) anterior superior portion connected with frontal, parietal, and temporal lobes; 3) anterior inferior portion connected with frontal and temporal lobes. These tractography-derived connectivity gradients are consistent with macaque tracer studies showing that insular subregions are embedded in distinct, somatotopically organized structural circuits with selective connections to frontal, parietal, and temporal areas^14^.

To go beyond structural connectivity, cortico-cortical evoked potentials (CCEPs) represent a form to assess in vivo brain effective connectivity, i.e. the effect one area has on another one. It is based on the application of low frequency (i.e., typically 1 Hz) electrical stimulation in patients undergoing stereo electroencephalography (SEEG) monitoring due to refractory epilepsy^15^. This method allows to measure the causal influence between brain regions since the stimulation at one site may result in an evoked response at distant sites, leading to a direct measure of effective connectivity. This approach has high level of temporal and spatial resolution and allows to measure directionality of connectivity because a stimulated brain region A can exert an evoked response at region B but the relationship may not be reciprocal^16^.

Effective connectivity of the human insula has been documented by only few studies with a relatively limited amount of data^9,17–20^. Dionisio et al.^9^ measured CCEPs in 30 participants, demonstrating that insula indeed connects with temporal, frontal, parietal, occipital, and limbic regions. Almashaikhi et al.^19^, with 11 participants, also revealed insular connectivity with perisylvian structures, pericentral cortex, temporal neocortex, mesial temporal structures, orbitofrontal and dorsolateral frontal cortices. Moreover, this study showed that insular effective connectivity differed across its gyri and partially diverged from patterns observed in nonhuman primates, supporting Afif et al.’s^20^ finding obtained on 25 patients and highlighting the need for more detailed investigation of the insula.

Due to the scarcity of published data, it remains unsolved how effective connectivity of the different insular subregions precisely targets the rest of the brain, or how it integrates incoming multilobar information. Fulfilling current literature limitations, this study aims to report the effective connectivity of human insula at a high-resolution with appropriate statistical power to serve as a reference, using the extensive multicenter CCEP database of the F-TRACT project^21,22^. To deal with SEEG subsampling issues, we mapped neurophysiological data on high-resolution anatomical atlases of the insula with intermediate resolution for the rest of the brain, an approach optimized to map brain sub-systems with high statistical power, which we recently proposed for the lateral frontal cortex^23^.

## Materials and methods

### SEEG acquisition and CCEP processing

The multicenter F-TRACT database (see consortium details in Appendix) comprises CCEPs induced by low-frequency (≤1Hz) bipolar electrical stimulations performed during SEEG explorations as in standard clinical practice for epilepsy surgery. All patients agreed and signed an informed consent for the re-use of their data for the F-TRACT protocol which was approved by the International Review Board at INSERM (protocol number: INSERM IRB 14-140), in accordance with the Declaration of Helsinki.

We selected the subjects with at least one electrode contact located in insula: from the 1223 curated patients in the dataset, it resulted in 897 (gender: 459 females, 435 males, 3 unknown; age: 26±14 years) from 21 epilepsy surgery centers. Extensive details on the anatomical and functional preprocessing steps to compute the CCEPs used for this study can be found in previous reports^21,24^ and in supplementary material (section 1).

SEEG was recorded using local clinical practice during low frequency (1 Hz or below) electrical stimulations between two contiguous contacts located either in the grey (56% whole-brain; 77% insula) or close to (<0.5 cm) white matter (42% whole-brain; 23% insula), using either monophasic 20% or biphasic 80% electrical pulses. The number of pulses in a stimulation run was 30±24. Stimulation parameters varied in pulse intensity (3±1 mA; 82% 3-4 mA) and duration (1±0 ms; 91% 0.5-1.05 ms). The patients were implanted with 120±36 electrode contacts in whole-brain and 6±6 electrode contacts in insula, being each contact neuroanatomically labeled after the co-registration of SEEG contacts with the pre-operative MRI^25^ and normalisation of contact coordinates to the MNI space used by insular anatomical atlases. To label a contact, a sphere of 1cm diameter around its MNI coordinates was considered and it was attached to the region of interest (ROI) with the largest number of voxels in it^25^. The resulting dataset for this study includes 70031 stimulations in the whole brain with recordings in insula and 4153 stimulations in insula with recordings in the whole brain. The type of epilepsy included are distributed as follows: temporal (24% left, 22% right), frontal (16% left, 14% right), insular (9% left, 8% right), parietal (7% left, 5% right), central (5% left, 4% right), occipital (3% left, 3% right), indeterminate (3%), and multifocal (6%). Stimulations were performed under no or various antiepileptics medication (230 patients without medication) and with the patients awake and at rest.

The whole dataset was fully curated, with automatic procedures and final supervised validation, to remove corrupted data (e.g. bad channels, ineffective current stimulation). For each recording contact and stimulation run, the evoked response averaged over stimulation pulses, i.e. the CCEP, was computed after artifact blanking, band-pass filtered [1,45] Hz to limit the effect of high frequencies potentially related to epileptogenic responses, and z-normalized using the prestimulus baseline period [-200,-10] ms^21^. It was considered significant if its absolute value reached the z-score threshold of 5 during [10,100] ms time window, having been this threshold value identified as an appropriate trade-off between sensitivity and specificity for subsequent group statistics^21^. In this study, we focused on a [10,100] ms time window, which encompasses both short-latency and intermediate-latency connections^16^. To verify that our results were not sensitive to the choice of the time window, we computed connectivity matrices using multiple window lengths and quantified their similarity. As shown in supplementary material (Section 2), connectivity matrices derived from different time windows were strongly correlated (r > 0.93), indicating strong consistency across window choices.

In the case of a significant response, the peak delay was computed for the first significant peak to infer feedforward connections^21^. Peak delay corresponded to the latency of the first peak above the z-score threshold from 10ms after the stimulation pulse.

### Group level analysis

Each brain connection is defined for a specific pair of stimulation and recording ROIs. Assuming that similar first CCEP components are recorded between individuals for a given connection^26^, the outcomes over the whole population of a particular brain connection were calculated over all recording contacts labeled in a particular ROI (recording brain region) due to all stimulation contacts in the same or another particular ROI (stimulation brain region). Each recording contact was considered responsive or not if it presented or not a significant response, respectively. Connectivity probability and peak delay were then calculated considering the aggregated responses from all contacts (and thus all participants) within a given connection.

The probabilistic connectivity for a connection was defined as the ratio between the number of recorded significant responses over the number of stimulations. Thus, connectivity probabilities varied between 0 (no response was significant) and 1 (all responses were significant), increasing proportionally to the number of significant responses. The peak delay was calculated over all significant responses for a given connection. Median was chosen to limit the impact of outliers in the group level analysis (a broader descriptive statistic is available in supplementary data, comprising average, standard deviation, quartile 25%, and quartile 75% of response amplitude and peak delay).

To guarantee the robustness of our results, we set a minimum of 50 available responses from at least 2 patients in each connection to compute the outcomes of effective connectivity. These values were selected according to a trade-off between brain coverage and confidence interval (CI), expecting to decrease confidence interval length and thus get more precise connectivity without losing considerable brain coverage for studying connectivity between the insula and the whole brain. The effect of these requirements on the outcome of probability can be observed in section 3 of supplementary material. Our results comprised a range of 85-90% brain coverage and 0.02-0.05 average CI length for probability. Moreover, surrogate data was used to define meaningful probabilities of connectivity by being above the surrogate probabilities (surrogate threshold). For a p-value=0.001, the surrogate probabilistic threshold was 0.15 considering a [0, 100] ms time window after events. In this manner, only probabilities above 0.15 were considered meaningful to describe effective connectivity between stimulated and recorded brain regions. More details can be found in section 4 of supplementary material. The median peak delay was calculated only for connections that met two criteria: at least 50 significant responses from a minimum of 2 patients, and a connectivity probability exceeding the surrogate threshold.

### Anatomical regions of interest

We used the “Lausanne2018,” “Montreal,” and “Jülich v3.0” atlases to compute the results for this study. Both the Montreal^12^ and Jülich^27^ atlases offer high-resolution subdivisions of the insula, comprising 19 and 16 parcels (i.e., brain regions), respectively. Since the Montreal atlas does not include whole-brain coverage, we used the Montreal and Jülich atlases to define insula regions, while the Lausanne2018^28^ was employed to whole-brain regions.

#### Insula atlas schemes

Insula’s Montreal atlas aligns with anatomical division of insula, derived from high angular resolution diffusion imaging tractography analysis conducted on 46 healthy young adults^12^, whereas the Jülich is a cytoarchitectonic atlas based on histological sections from 23 postmortem brains^27^, which aligns with architectonically division of insula. The Montreal atlas was selected for the main text results to facilitate the comparison with MR DTI results published with this atlas^12^. The inclusion of the Jülich atlas allows us to assess the consistency of our findings by correlating them with a classical cytoarchitectonic decomposition.

In this study, we employed three levels of insular resolution —low, mid, and high— for both Montreal and Jülich atlases. Specifically: a) low resolution consists of a single insula parcel obtained by merging all insular parcels within the scheme (Montreal1 and Jülich1); b) mid resolution considers four insula parcels, closely aligned with the widely adopted anterior-posterior and superior-inferior anatomical and cytoarchitectonic divisions (Montreal4 and Jülich4); c) high resolution consists of full raw atlas resolution (Montreal19 and Jülich16). Additionally, a mid-to-high resolution is included in supplementary data comprising seven and ten parcels for Montreal and Jülich. These were derived as optimal trade-offs between spatial resolution and the distribution density of F-TRACT data while preserving the underlying sulco-gyral anatomy^5,12,29^ and cytoarchitectonic division^1^, respectively. Details on the criteria used to define these resolution levels are provided in supplementary material (section 5.1).

#### Whole brain atlas scheme

Lausanne2018 atlas is based on anatomical landmarks derived from diffusion spectrum imaging in a cohort of 40 healthy individuals^30^. Through iterative subdivision of each ROI, the atlas is available at multiple resolutions—scales 1, 2, 3, and 4—corresponding, respectively, to 94, 141, 242, and 472 parcels for right and left symmetric hemispheres^28^. For this study, we considered the scale1 (supplementary data), scale2 (used in the main text), and scale3 resolutions (supplementary data), selected as optimal trade-offs between spatial resolution and the distribution density of data across brain regions. Regions were grouped into the following six main systems: frontal, central, parietal, temporal, limbic, and occipital. The limbic system includes amygdala, hippocampus, parahippocampal, temporal pole, entorhinal, anterior cingulate, and orbitofrontal cortex^31^. Description of Lausanne-scale2’s parcels and selected systems can be found in section 5.2 of supplementary material.

### Connectivity analysis

Connectivity probability (0-1) and peak delay (ms) matrices were generated for Montreal1 stimulations and Lausanne-scale2 recordings to describe efferent connectivity between insula and the whole brain. Brain maps were created from connectivity matrices for an intuitive representation of connectivity in the medial and lateral views of ipsilateral and contralateral brain hemispheres. Efferent probabilities were also computed for Montreal4 and Montreal19 stimulation to assess mid- and high-resolution insular effective connectivity with the whole brain, respectively.

Afferent probability was explored by generating a probability matrix for Lausanne-scale2 stimulations and Montreal1, Montreal4, or Montreal19 recordings. While efferent (outgoing) connectivity represents responses in the whole brain when insula is stimulated, the afferent connectivity (incoming) represents responses in insula when stimulation is elsewhere in the brain. A measure of directionality was computed by subtracting the efferent and afferent probabilities corresponding to the same connection and then normalized by the maximum probability (between efferent and afferent probabilities). In this manner, directionality varied between −1 and 1. Zero directionality meant that connectivity was equal in both directions (from insula to the whole brain and from the whole brain to insula), −1 meant that insula connectivity was purely afferent, and 1 meant insula connectivity was purely efferent. All the results were calculated for Jülich atlas similarly to Montreal (supplementary material).

Whole-brain analyses showed high degree of symmetry between left and right hemispheres (r=0.80; strong correlation was considered for *r* > 0.7^32^). Therefore, for this report, we assumed interhemispheric differences were small and, to increase statistical power and spatial coverage for the highest resolution where insular subregions are tiny, we merged the responses between right and left hemispheres for ipsi (arbitrarily represented on the left side) and contralateral (arbitrarily represented on the right side) connectivity.

Interested readers can refer to supplementary materials (Figure 14 from supplementary material) to access to left and right connectivity. Additionally, we provided efferent and afferent probability brain maps for Montreal7 with Lausanne-scale2, Montreal4 with Lausanne-scale3, and Montreal19 with Lausanne-scale1. Matrices of the total number of responses and patients were also included.

### Statistical analysis

A z-test for proportions based on binomial distribution with a significance level of 0.05 was employed to statistically compare the observed probabilities of connectivity. The following null hypotheses were tested: (a) there are no statistically significant differences between efferent and afferent probabilities. Directionality is measured in case of a significant difference found; (b) there are no statistically significant differences in how the 19 insular subregions connect with the rest of the brain. The strongest insular connection (highest probability of connectivity) to a given brain region was compared with the remaining 18 insular connections. A difference was detected if at least two insular connections showed a statistical difference. The insular subregion with the strongest connection, along with any insular subregions with connections statistically similar to it, were considered the predominant insular contributors of connectivity to the given brain region. For all hypotheses, the Benjamini–Hochberg procedure was applied to control the false discovery rate due to multiple comparisons. Supplementary, effect size was calculated using Cohen’s h, defined as the absolute difference between the arcsine–square-root–transformed probabilities of connectivity being compared^33^.

Whole brain and insula predominance maps were computed according to the results from hypothesis (b). Whole brain predominance map was created to identify the insula subregion stimulated with highest probability of connectivity to each brain region if a statistical difference was found between efferent probabilities of high-resolution insular connections. Insula predominance maps show the percentage of predominant insular connections in each brain system (defined as frontal, parietal, temporal and occipital lobes, central regions, and limbic regions). They were created for efferent and afferent connections.

Additionally, Spearman correlation was computed between probability matrices as follows: (a) Montreal vs Jülich stimulation to quantify the consistency between the atlas schemes; (b) all CCEPs vs only CCEPs recorded with no epileptiform spikes, detected with Delphos software ^34^, to quantify the influence of underlying epileptic activity; and (c) both grey and white matter contacts vs only grey matter contacts to quantify the influence of grey vs. white matter stimulation. These correlation analyses used probability matrices from Montreal1 (or Julich1) stimulations and Lausanne-scale2 recordings. Peak delay was also tested for correlation with probability. Strong correlations were considered for correlation coefficient *r* > 0.7^32^.

## Data availability

Raw data cannot be distributed because of personal data protection sensitive issues. Fully processed data at the group level are available for download in the following GitHub link: https://github.com/ins-amu/ft-insula

## Results

### Connectivity of whole insula

Figures 1A and 1B show the number of recordings and efferent probability of connectivity of whole insula, respectively. Significant probabilities ranged between 0.15 and 0.73 englobing ipsilateral frontal, parietal, central, temporal, and limbic systems. Ipsilateral occipital probabilities did not pass the surrogate threshold. Contralateral regions were less recorded in comparison with ipsilateral hemisphere and showed lower connectivity probabilities, being mostly under the surrogate threshold.

**Figure 1.**
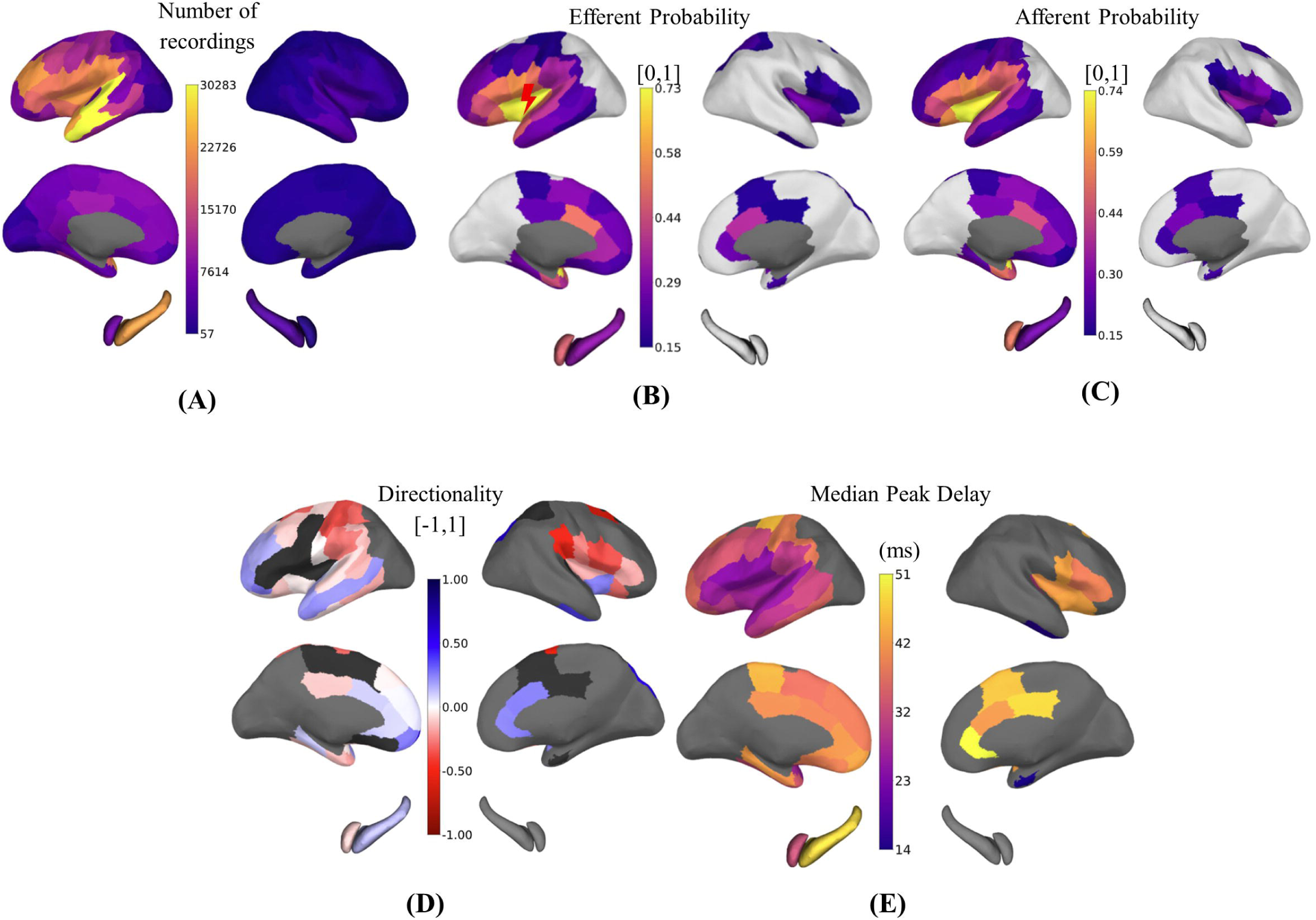
Connectivity of whole insula (Montreall). **(A)** Number of recordings; **(B)** Efferent connectivity probability; **(C)** Afferent connectivity probability; **(D)** Directionality; **(E)** Median peak delay (ms). Light grey areas in (B-C) represent probabilities below surrogate threshold. No color appears if the requirement for minimum number of responses is not met. Dark grey color in (D) represents no statistically significant differences between efferent and afferent probabilities (p-value > 0.05).

Considering the insula being stimulated, the probability of connectivity decreased as the distance from insula increased. Mid-to-high probabilities (0.44-0.73; Table 1) were found in ipsilateral insula; frontal (pars triangularis and pars opercularis); central (precentral and postcentral); temporal (transverse temporal and superior temporal); and limbic systems (lateral orbitofrontal, caudal anterior cingulate, entorhinal, and amygdala).

**Table 1.** Summary of effective connectivity of whole insula. Identification of brain regions per system (frontal, parietal, central, temporal, limbic, and occipital) which revealed mid-to-high insular probabilistic connectivity (0.44-0.73), efferent and afferent directions of connectivity (−0.60:0.71), and low-to-mid median peak delays (14-32 ms) considering Montreal1 stimulations and Lausanne2018-scale2 recordings

| **SYSTEM** | **MID-TO-HIGH**<br>**EFFERENT**<br>**PROBABILITIES**<br>**(0.44-0.73)** | **EFFERENT**<br>**DIRECTION**<br>**[0.00:0.71]** | **AFFERENT**<br>**DIRECTION**<br>**[-0.60:0.00]** | **LOW-TO-MID**<br>**MEDIAN PEAK**<br>**DELAY**<br>**(14-32 MS)** |
| --- | --- | --- | --- | --- |
| **FRONTAL** | Pars triangularis<br>Pars opercularis | Pars orbitalis<br>Frontal pole<br>Superior frontal<br>Middle frontal | Superior frontal<br>Middle frontal | Pars triangularis<br>Pars opercularis<br>Pars orbitalis<br>Middle frontal |
| **PARIETAL** | None | None | Supramarginal<br>Superior parietal<br>Posterior cingulate | Supramarginal |
| **CENTRAL** | Precentral<br>Postcentral | None | Precentral<br>Postcentral | Precentral<br>Postcentral |
| **TEMPORAL** | Transverse temporal<br>Superior temporal | Superior temporal<br>Middle temporal | Superior temporal<br>Banks<br>Inferior temporal<br>Fusiform | Transverse temporal<br>Superior temporal<br>Middle temporal<br>Banks<br>Fusiform |
| **LIMBIC** | Orbitofrontal<br>Anterior cingulate<br>Entorhinal<br>Amygdala | Orbitofrontal<br>Anterior cingulate<br>Parahippocampal<br>Hippocampus | Orbitofrontal<br>Entorhinal<br>Temporal pole<br>Amygdala | Orbitofrontal<br>Temporal pole<br>Amygdala |
| **OCCIPITAL** | None | None | None | None |

Considering the whole brain being stimulated, mid-to-high afferent probabilities (0.44-0.74; Figure 1C) were found in ipsilateral insula; frontal (pars triangularis and pars opercularis); central (precentral and postcentral); transverse temporal; supramarginal; and limbic regions (lateral orbitofrontal, entorhinal, temporal pole, and amygdala).

Figure 1D shows the directionality of probabilistic effective connectivity between insula and the whole brain, varying between −0.60 and 0.71. The pattern of connectivity is summarized in Table 1. Briefly, predominant efferent directionality was found between insula and ipsilateral frontal, temporal and anterior cingulate regions. Contralaterally, insula mostly signalled to superior frontal, inferior temporal, and limbic system. Predominant afferent directionality was noticed between insula and ipsilateral frontal, parietal, central, temporal and limbic regions. Contralaterally, insula mostly received inputs from frontal, central and orbitofrontal regions.

Figure 1E shows the median peak delay of connectivity between insula and the whole brain. The values of peak delay ranged from 14 to 51 ms and demonstrated a pattern negatively correlated with probability (r=-0.35 and p-value=0.0004). The fastest responses (14-32 ms; Table 1) occurred in ipsilateral insula and in surrounding areas.

Connectivity maps were also generated with the Jülich atlas scheme (section 7 of supplementary material). They were extremely similar (r ≥ 0.94), indicating little dependence to detailed contouring of insular parcels.

### Connectivity of insular subregions

Figure 2 shows the efferent probabilistic connectivity and directionality maps of insula at mid and high-resolution. Briefly, patterns of connectivity vary between superior-inferior and posterior-anterior insula axes.

**Figure 2.**
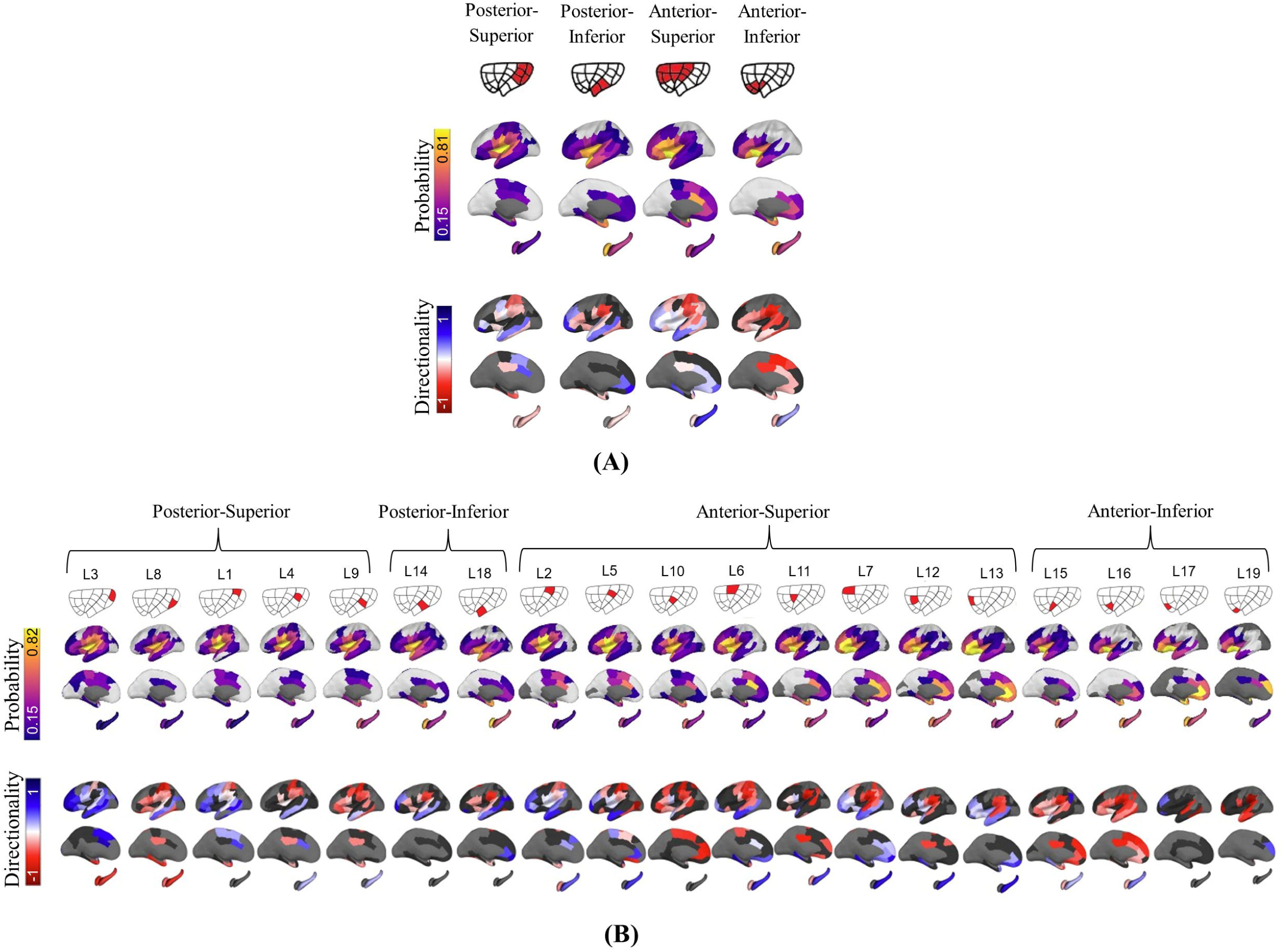
Insula connectivity at mid (A) and high (B) resolution. Efferent probabilistic connectivity and directionality of effective connectivity for **(A)** Montreal4 stimulations and **(B)** Montreall 9 stimulations. Light grey color represents probabilities below surrogate threshold. No color appears when the requirement for minimum number of responses is not met. Black color represents no statistically significant differences between efferent and afferent probabilities (p-value > 0.05).

Posterior superior insula is mainly connected with parietal, central, and temporal systems. Connections move from parietal to central systems along a caudal-to-rostral axis, and from central and parietal to temporal systems along a dorsal-to-ventral axis. Regarding directionality, posterior superior insula mostly receives input [-0.74:-0.05] from supramarginal, superior parietal, posterior cingulate, and inferior temporal and sends output [0.28] to the middle temporal.

Posterior inferior insula is primarily connected with temporal and limbic systems, moving from temporal to limbic along a caudal-to-rostral axis. It receives input [-0.80:-0.10] from inferior temporal, fusiform, entorhinal, and hippocampus; and sends output [0.06:0.56] to the middle temporal and rostral anterior cingulate.

Anterior superior insula connects with frontal, central, parietal, and limbic systems. Connections move from frontal, central, and parietal systems to limbic system along a dorsal-to-ventral axis and from parietal and central to frontal along a caudal-to-rostral axis. It receives input [-0.67:-0.07] from caudal middle frontal, supramarginal, superior and inferior parietal, posterior cingulate, postcentral, lateral orbitofrontal, temporal pole, and amygdala; and sends output [0.02:0.74] to rostral middle frontal, pars triangularis, anterior cingulate, medial orbitofrontal, parahippocampal, and hippocampus.

Anterior inferior insula is primarily connected with frontal, temporal, and limbic systems, moving from temporal system to frontal and limbic systems along a caudal-to-rostral axis. It receives input [-0.86:-0.04] from superior frontal, pars triangularis, pars opercularis, inferior temporal, cingulate, entorhinal, temporal pole, and amygdala; and sends output [0.49] to hippocampus.

### Predominance maps

Predominance maps (Figure 3 and Table 2) were finally generated to summarise information.

**Figure 3.**
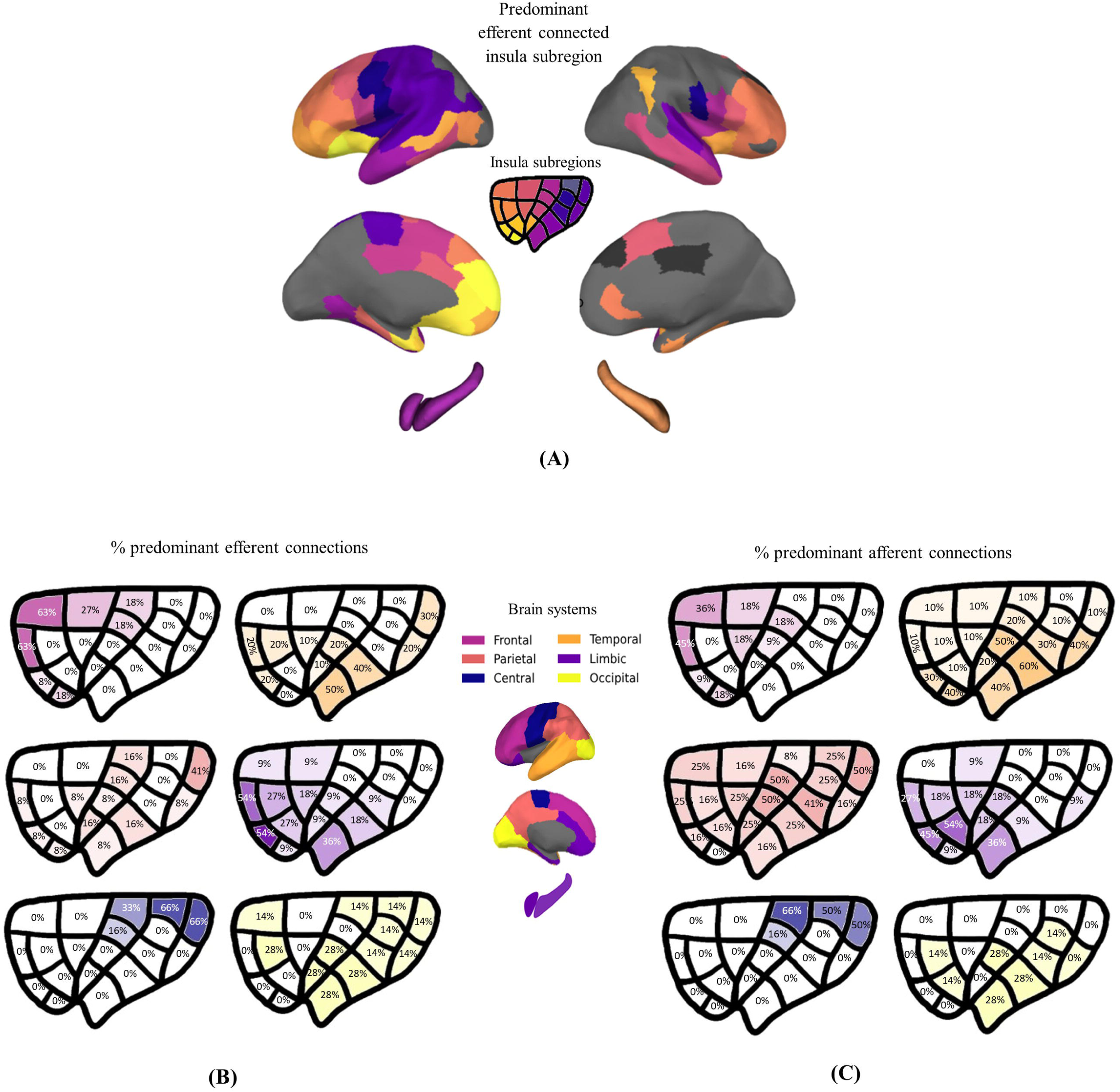
Predominance maps. **(A)** Brain map identifying predominant efferent connected insula subregions (1-19); **(B)** Insula efferent predominance map; **(C)** Insula afferent predominance map. Insula maps represent the percentage of predominant insular connections in the main brain system (frontal, parietal, central, temporal, limbic, and occipital). Black color in (A) means that all insula subregions are similarly connected to the given brain region (p-value > 0.05).

**Table 2.**
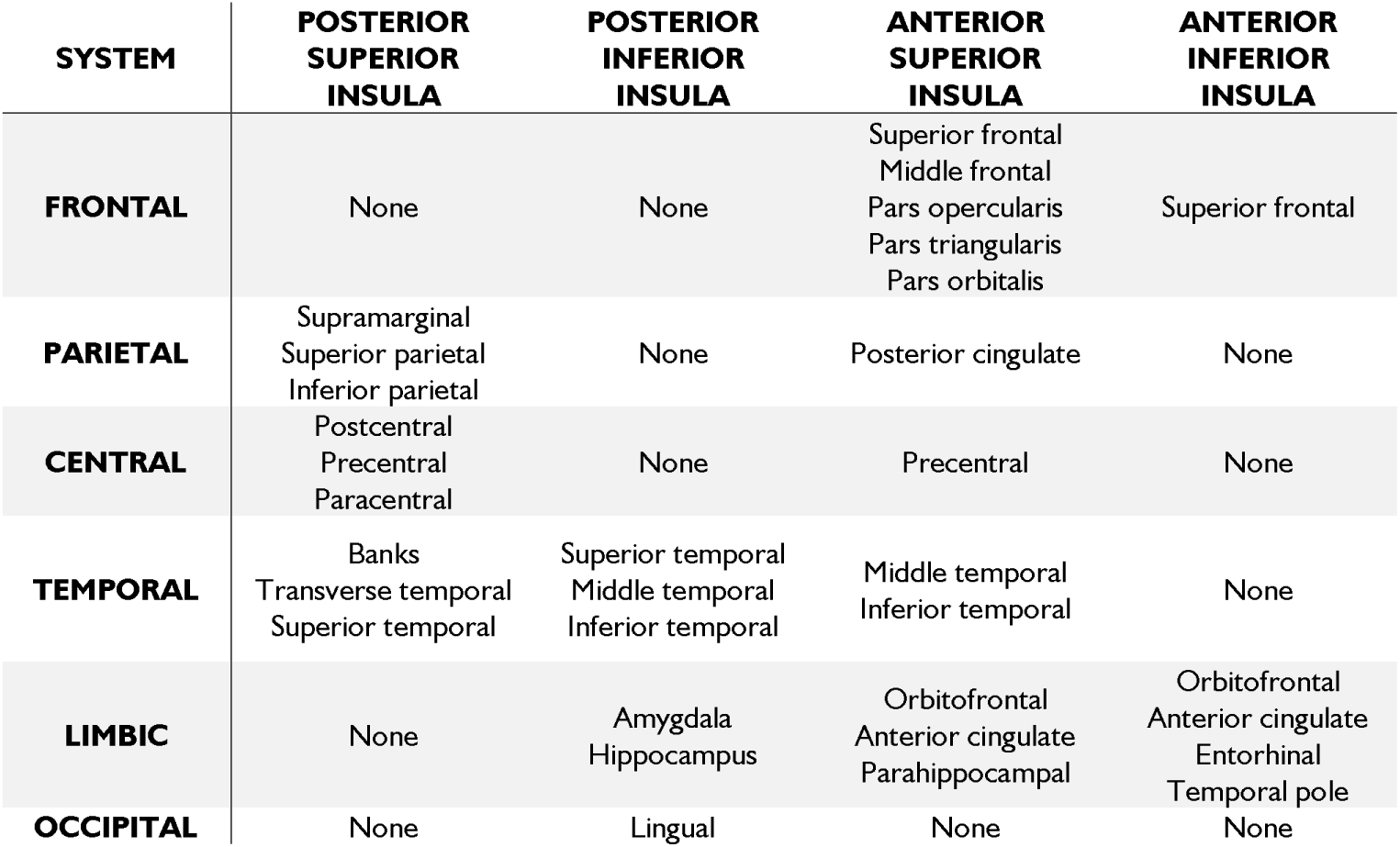
Insula predominance of effective connectivity per brain system. Identification of the highest efferent connected insula subregion (Montreal4 insula subregions: posterior superior, posterior inferior, anterior superior, and anterior inferior) to each brain region per system (frontal, parietal, central, temporal, limbic, and occipital).

Figure 3A shows the predominant efferent connected insula subregion to each region of the cortex. Posterior superior insula is predominantly connected to ipsilateral parietal (supramarginal, superior and inferior parietal), central (postcentral, precentral, and paracentral), and temporal systems (banks, transverse and superior temporal). Posterior inferior insula is predominantly connected to ipsilateral temporal system (superior, middle, inferior temporal regions), amygdala, hippocampus, and lingual regions. Anterior superior insula is predominantly connected to ipsilateral frontal system (superior and middle frontal, pars opercularis, pars triangularis, and pars orbitalis), posterior cingulate, precentral, temporal cortex (middle and inferior temporal), and limbic system (medial orbitofrontal, caudal anterior cingulate, parahippocampal). Finally, anterior inferior insula is predominantly connected to ipsilateral superior frontal cortex and limbic system (orbitofrontal, rostral anterior cingulate, entorhinal, and temporal pole).

Contralaterally, we see predominant probabilistic connectivity between posterior superior insula with postcentral area and temporal regions (transverse and superior temporal); anterior superior insula with frontal regions (superior frontal, pars triangularis, pars opercularis, and rostral middle frontal), precentral area, middle temporal, and limbic regions (entorhinal and hippocampus); and anterior inferior insula with inferior parietal. The values of effect size varied between 0.36-1.81 (Figure 8 of supplementary material).

To refine the previous analysis, the percentage of predominant efferent (Figure 3B) and afferent (Figure 3C) insular connections to the main brain systems were then mapped for each insula subregion. Most insular subregions show a degree of connection with multiple brain systems, organized topologically. Frontal system is predominantly connected with anterior superior insula with decreasing percentage of predominant connectivity along dorsal-ventral and rostral-caudal axis (8-63%, cumulative 215%). Parietal and central systems are predominantly connected to posterior superior insula with decreasing predominance along the dorsal-ventral and caudal-rostral axis (8-41% parietal, cumulative 161%; 16-66% central, cumulative 181%). Temporal and occipital systems are predominantly connected to posterior inferior insula with decreasing predominance along ventral-dorsal axis (10-50% temporal, cumulative 240%; 14-28% occipital, cumulative 238%). The limbic system is predominantly connected to anterior inferior insula with decreasing predominance along a ventral-dorsal and rostral-caudal axis (9-54%, cumulative 288%).

By comparing efferent and afferent predominance maps, we see that frontal (9-45%, cumulative 171%), limbic (9-54%, cumulative 234%), and occipital systems (14-28%, cumulative 154%) present lower insula predominance in the afferent maps; while parietal (16-50%, cumulative 462%) and temporal (10-60%, cumulative 420%) present higher insula predominance in the afferent maps.

### Control analyses

Additional analyses were done to assess the robustness of our results. Firstly, we compared probability matrices using all channels vs. considering only channels with no interictal spikes (Figure 12 from supplementary material), which demonstrated a correlation coefficient of r=0.93 (p-value=3.8e-49). This result suggests limited influence of epilepsy in the inferred pattern of insular connectivity, which has probably been smeared out by the large statistics and the various epilepsy types. Secondly, we compared probability matrices using both grey and white matter labelled contacts vs using only grey matter labelled contacts (Figure 13 from supplementary material) resulting in a correlation coefficient of r=0.97 (p-value=1.1e-72). These results suggest similar activation of neural pathways whether stimulation contacts were within or close to the grey matter.

## Discussion

### Patterns of insula effective connectivity

In line with previous CCEP studies of the insula^9,19^, we found insular connectivity with pars opercularis, supramarginal gyrus, temporal (primary auditory cortex and lateral superior temporal gyrus), and limbic systems (orbitofrontal and amygdala). As Dionisio et al.^9^, we found connectivity between insula and pars triangularis and anterior cingulate. As Almashaikhi et al.^19^, we found connectivity between insula and primary motor and somatosensory cortices (precentral and postcentral, respectively) and entorhinal. Using the parcellation scheme of Montreal, structural connectivity analyses from Ghaziri et al.^12^ based on MR diffusion tractography also revealed insula connection with frontal system (pars triangularis, pars orbitalis, pars opercularis, superior frontal), supramarginal, central (precentral and postcentral), temporal (superior temporal and transverse temporal), and limbic systems (orbitofrontal, entorhinal, temporal pole, and anterior cingulate). As in Ghaziri et al.^12^, we also found connections with superior parietal, posterior cingulate, precuneus, fusiform, parahippocampal, and occipital system (cuneus, lingual, and lateral occipital), but with low probability (<0.3). In comparison to Ghaziri et al.^12^, our study brings as new information the insular connection with amygdala and hippocampus that were not documented.

Regarding the topological organization of insular connectivity, in accordance with Ghaziri et al.^12^, we demonstrated posterior superior insula connectivity with parietal and temporal lobes; posterior inferior insula with temporal and occipital lobe; anterior superior insula with frontal, parietal, and temporal lobes; and anterior inferior insula with frontal and temporal lobes. However, Ghaziri et al.^12^ additionally showed posterior insula connectivity with frontal lobe that we did not identify. In fact, our results are consistent with models of interoceptive information processing presented in Evrard et al.^35^, in which posterior insular regions primarily interact with sensory areas, while anterior subdivisions show progressively stronger coupling with cognitive-affective areas.

Compared to previous studies, we bring significant information regarding the dynamics and directionality of insular connectivity.

Peak delays ranged from 14 to 51 ms, with the fastest responses (14–32 ms) occurring in the ipsilateral insula and nearby regions. In line with Lemarechal et al.^24^, primary areas, namely, primary somatosensory, primary motor cortex, and primary auditory cortex were the ones exhibiting fastest responses.

Overall, the insula predominantly received afferent inputs from widespread parietal, central, temporal, frontal, and limbic regions, while sending efferent outputs mainly to frontal, temporal, cingulate, and hippocampal regions. In accordance with our results of directionality of connectivity, Bastuji et al.^36^ and Allen et al.^37^ reported that insula integrates limbic with sensory inputs to initiate a perceptual decision on the stimulus painfulness and control signals arising in the prefrontal and cingulate cortex^38^, causally regulating the interaction between sensory and attentional networks^39–41^. Moreover, Huang et al.^42^ observed asymmetry in insula-hippocampus communication suggesting the hippocampus works as a coordinator, integrating and responding to inputs from specialized cortical areas such as insula.

### Functional interpretation

Sensorimotor integration is supported in our study by insular connection with paracentral, postcentral, supramarginal gyrus, and superior temporal sulcus^7,13,43–45^. Insular connections with superior temporal and supramarginal regions relate auditory processing to motor adaptation for speech^46,47^, while postcentral connectivity suggests roles in touch, proprioception, and body awareness^9^. Connectivity with superior temporal, supramarginal, and postcentral regions also aligns with the insula’s vestibular contributions^9^. Besides the insula role on sensorimotor integration, our results on insula-lingual connections also support insula role in visual consciousness^10,48^.

Our findings on insula–orbitofrontal connectivity support decision-making role in which insula inputs contribute to orbitofrontal hedonic encoding^49,50^, and to cognitive appraisal of reward and punishment in social contexts^7,13,43,44^. Moreover, our findings on insula connectivity with Von Economo neuronal network (amygdala, entorhinal cortex, orbitofrontal cortex, and anterior cingulate) suggest insula role on emotional salience and homeostasis^51–54^

Prior studies claim for insula role in language^24,55–57^, reflected here in its connectivity with lateral inferior frontal area and superior temporal areas, which support integration of speech production with auditory processing^7,45,58^. We also identified insula connections with pars triangularis, pars opercularis, and supramarginal gyrus which suggest insula involvement with the ventral attention network^59^, while identified connections with rostral middle frontal gyrus reflect involvement in goal-directed attention within the dorsal attention network^1360^.

At higher resolution, we identified posterior insula connections with parietal, central, and temporal regions and anterior insula connections with limbic and frontal systems, which align with literature by mapping posterior insula to sensorimotor function and anterior insula to social-emotional and cognitive functions^61^. Cecchi et al.^62^ found that direct electrical stimulation had dissociable effects on choice behavior on a dorso-ventral axis in the anterior insula which aligns with our identified anterior insular connectivity with orbitofrontal and prefrontal cortex. Additionally, Simone et al.^14^ indicated that posterior insula integrates vestibular, auditory, and somatosensory inputs while anterior insula supports socio-emotional processes and reward evaluation.

### Limitations

These findings should be interpreted considering the following limitations. First, mechanisms underlying CCEPs remain debated. Although electrical stimulation is generally assumed to activate pyramidal neurons and propagate orthodromically, stimulation of axons in white matter may produce antidromic propagation, which cannot be excluded^15,63,64^. We retained white matter contacts (23%) for statistical robustness. However, analyses restricted to grey matter contacts yielded highly correlated patterns (r = 0.97). This suggests that direct electrical stimulation may activate predominantly the axons of pyramidal cells, explaining why there is little difference between white (close to grey) and grey matter stimulation connectivity patterns^65^.

Second, despite strong correlations between stimulation–recording schemes (r ≥ 0.94), the inherent sampling bias in electrode placement (driven by clinical requirements) may still cause underrepresentation of certain regions, leading to false negatives, as illustrated by limited insula–occipital connections.

Third, statistical analyses assumed measurement independence in binomial proportion tests, although recordings may share dependencies (due to factors such as clinical center, stimulation waveform, pulse parameters, and number of contacts per region). Nevertheless, the large and diverse cohort, and consistency of results with prior literature suggest minimal impact.

Fourth, although the influence of epilepsy on CCEPs remains debated, our dataset included a large and heterogeneous cohort, with many contacts located in non-epileptic tissue, which should reduce pathological bias. Some studies report no alteration of the early component in hyper-excitable zones^66^, while others note only high-frequency changes^67^. Functional connectivity alterations seem restricted to the epileptogenic zone^68^, and 1Hz stimulation studies report no differences between epileptogenic and non-epileptogenic hemispheres^69^. Nonetheless, pathology-related effects cannot be entirely excluded, but control analyses using only zero-spike-count CCEPs yielded results strongly correlated with the full dataset (r = 0.93).

### Relevance to clinical applications

The present study, showing whole-brain effective connectivity mapping of the insula, has direct translational value. In epilepsy, seizures can start from the insula region and mimic temporal or frontal lobe seizures through its propagation patterns^70^, making accurate network characterization critical for implantation of intracerebral electrodes when needed. Beyond epilepsy, the insula is increasingly recognized as a transdiagnostic hub implicated in psychiatric and neurological disorders, including anxiety^71^, schizophrenia^72^, psychopathy^73^, anorexia nervosa^74^, Huntington’s disease, multiple sclerosis^75^, Alzheimer’s disease^76^, autism spectrum disorder^77^, and addiction^78^. By delineating the insula’s connectivity profile, our findings provide a network-level framework to inform diagnosis, guide targeted interventions, and support mechanistic research across a wide spectrum of brain disorders.

## Conclusion

This study provides a detailed mapping of insular effective connectivity. Insular connectivity was observed primarily in ipsilateral frontal system (pars triangularis, pars opercularis, pars orbitalis, superior frontal), supramarginal, central (postcentral, precentral), temporal (transverse temporal, superior temporal), and limbic systems (lateral orbitofrontal, anterior cingulate, entorhinal, temporal pole, amygdala, and hippocampus). Peak delays ranged from 14 to 51 ms, with the fastest responses in the insula surrounding areas. A gradient of connectivity is observed along both posterior–anterior and superior–inferior axes: posterior insula is mainly connected with parietal, central, temporal, and limbic regions, while the anterior insula additionally connects with frontal regions; superior insula is mainly connected with frontal, parietal, central, temporal, and limbic regions, whereas the inferior insula is primarily connected with temporal and limbic regions. Some regions exhibit directional preferences of connectivity. Insula seems to receive mostly from caudal middle frontal, parietal, primary somatosensory cortex, and amygdala, and to project to rostral middle frontal and hippocampus. Overall, these findings underscore the insula’s role as a convergence hub for interoceptive, exteroceptive, and cognitive-emotional information, providing a network-level framework to inform diagnosis, guide interventions, and advance mechanistic understanding across neurological and psychiatric disorders.

## Acknowledgements

We acknowledge the work of the following people, involved in the past in preparation of the F-TRACT dataset: François Tadel, Lena Trebaul, Pierre Deman, Viateur Tuyisenge, Jean-Didier Lemarechal, Carole Saubat, Raouf Zouglech, Maëlle Gueguen, Gina Catalina Reyes Mejia, Leila Ayoubian, Etienne Hugues.

## Funding

The research leading to these results has received funding from the European Research Council under the European Union’s Seventh Framework Programme (FP/2007–2013)/ERC Grant Agreement no. 616268 F-TRACT, the European Union’s Horizon 2020 Framework Programme for Research and Innovation under Specific Grant Agreement No. 785907 and 945539 (Human Brain Project SGA2 and SGA3), from the Agence Nationale de la Recherche (grant numbers ANR-21-NEUC-0005–01, ANR-22-CE17-0057-03 and ANR-22-PESN-0012), from the Swiss National Science Foundation (Sinergia 209470), and the National Institutes of Health grant number R01MH129018.

## Competing interests

The authors declare that they have no conflict of interest.

## Supplementary material

Supplementary material is available at Brain online.

## Appendix

F-TRACT consortium members: Claude Adam, Vincent Navarro, Arnaud Biraben, Anca Nica, Dominique Menard, Milan Brazdil, Robert Kuba, Jitka Kocvarová, Martin Pail, Irena Dolezalová, François Dubeau, Jean Gotman, Philippe Ryvlin, Jean Isnard, Hélène Catenoix, Alexandra Montavont, Sylvain Rheims, Fabrice Bartolomei, Agnès Trebuchon, Aileen McGonigal, Wenjing Zhou, Haixiang Wang, Sinclair Liu, Zhang Wei, Zhu Dan, Guo Qiang, Hu Xiangshu, Li Hua, Hua Gang, Wang Wensheng, Mei Xi, Feng Yigang, Rima Nabbout, Marie Bourgeois, Anna Kaminska, Thomas Blauwblomme, Mercedes Garces, Antonio Valentin, Rinki Singh, Liisa Metsahonkala, Eija Gaily, Leena Lauronen, Maria Peltola, Francine Chassoux, Elizabeth Landre, Philippe Derambure, William Szurhaj, Maxime Chochois, Edouard Hirsch, Maria Paola Valenti, Julia Scholly, Luc Valton, Marie Denuelle, Jonathan Curot, Rodrigo

Rocamora, Alessandro Principe, Miguel Ley, Ioana Mindruta, Andrei Barborica, Stefano Francione, Roberto Mai, Lino Nobili, Ivana Sartori, Laura Tassi, Louis Maillard, Jean-Pierre Vignal, Jacques Jonas, Louise Tyvaert, Mathilde Chipaux, Delphine Taussig, Philippe Kahane, Lorella Minotti, Anne-Sophie Job, Veronique Michel, Marie de Montaudoin, Jerôme Aupy, Viviane Bouilleret, Ana Maria Petrescu, Pascal Masnou, Claire Dussaule, Marion Quirins, Delphine Taussig, Carmen Barba, Renzo Guerrini, Matteo Lenge and Elisa Nacci.

## Notes

### Competing Interest Statement

The authors have declared no competing interest.

### Author Declarations

Ethics committee/IRB of Institut National de la Sante et de la Recherche Medicale (protocol number: INSERM IRB 14-140) gave ethical approval for this work

## References

1. Quabs J, Caspers S, Schöne C, et al. Cytoarchitecture, probability maps and segregation of the human insula. Neuroimage. 2022;260(January):119453. doi:10.1016/j.neuroimage.2022.119453

2. Nieuwenhuys R. The insular cortex. In: ; 2012:123-163. doi:10.1016/B978-0-444-53860-4.00007-6

3. AUGUSTINE J. Circuitry and functional aspects of the insular lobe in primates including humans. Brain Res Rev. 1996;22(3):229–244. doi:10.1016/S0165-0173(96)00011-2

4. Tanriover N, Rhoton AL, Kawashima M, Ulm AJ, Yasuda A. Microsurgical anatomy of the insula and the sylvian fissure. J Neurosurg. 2004;100(5):891–922. doi:10.3171/jns.2004.100.5.0891

5. Türe U, Yaşargil DCH, Al-Mefty O, Yaşargil MG. Topographic anatomy of the insular region. J Neurosurg. 1999;90(4):720–733. doi:10.3171/jns.1999.90.4.0720

6. Türe U, Yaşargil MG, Al-Mefty O, Yaşargil DCH. Arteries of the insula. J Neurosurg. 2000;92(4):676–687. doi:10.3171/jns.2000.92.4.0676

7. Cauda F, D’Agata F, Sacco K, Duca S, Geminiani G, Vercelli A. Functional connectivity of the insula in the resting brain. Neuroimage. 2011;55(1):8–23. doi:10.1016/j.neuroimage.2010.11.049

8. Mesulam M Marse., Mufson EJ. Insula of the old world monkey. III: Efferent cortical output and comments on function. J Comp Neurol. 1982;212(1):38–52. doi:10.1002/cne.902120104

9. Dionisio S, Mayoglou L, Cho SM, et al. Connectivity of the human insula: A cortico-cortical evoked potential (CCEP) study. Cortex. 2019;120:419–442. doi:10.1016/j.cortex.2019.05.019

10. Bisenius S, Trapp S, Neumann J, Schroeter ML. Identifying neural correlates of visual consciousness with ALE meta-analyses. Neuroimage. 2015;122:177–187. doi:10.1016/j.neuroimage.2015.07.070

11. Friston KJ. Functional and Effective Connectivity: A Review. Brain Connect. 2011;1(1):13–36. doi:10.1089/brain.2011.0008

12. Ghaziri J, Tucholka A, Girard G, et al. The Corticocortical Structural Connectivity of the Human Insula. Cereb Cortex. 2017;27(2):1216–1228. doi:10.1093/cercor/bhv308

13. Cloutman LL, Binney RJ, Drakesmith M, Parker GJM, Lambon Ralph MA. The variation of function across the human insula mirrors its patterns of structural connectivity: Evidence from in vivo probabilistic tractography. Neuroimage. 2012;59(4):3514–3521. doi:10.1016/j.neuroimage.2011.11.016

14. Simone L, Caruana F, Elena B, et al. Anatomo-functional organization of insular networks: From sensory integration to behavioral control. Prog Neurobiol. 2025;247(October 2024):102748. doi:10.1016/j.pneurobio.2025.102748

15. Matsumoto R, Nair DR, LaPresto E, et al. Functional connectivity in the human language system: a cortico-cortical evoked potential study. Brain. 2004;127(10):2316–2330. doi:10.1093/brain/awh246

16. Matsumoto R, Nair DR, LaPresto E, Bingaman W, Shibasaki H, Lüders HO. Functional connectivity in human cortical motor system: A cortico-cortical evoked potential study. Brain. 2007;130(1):181–197. doi:10.1093/brain/awl257

17. Lacuey N, Zonjy B, Kahriman ES, et al. Homotopic reciprocal functional connectivity between anterior human insulae. Brain Struct Funct. 2016;221(5):2695–2701. doi:10.1007/s00429-015-1065-0

18. Almashaikhi T, Rheims S, Ostrowsky-Coste K, et al. Intrainsular functional connectivity in human. Hum Brain Mapp. 2014;35(6):2779–2788. doi:10.1002/hbm.22366

19. Almashaikhi T, Rheims S, Jung J, et al. Functional connectivity of insular efferences. Hum Brain Mapp. 2014;35(10):5279–5294. doi:10.1002/hbm.22549

20. Afif A, Minotti L, Kahane P, Hoffmann D. Anatomofunctional organization of the insular cortex: A study using intracerebral electrical stimulation in epileptic patients. Epilepsia. 2010;51(11):2305–2315. doi:10.1111/j.1528-1167.2010.02755.x

21. Trebaul L, Deman P, Tuyisenge V, et al. Probabilistic functional tractography of the human cortex revisited. Neuroimage. 2018;181(June):414–429. doi:10.1016/j.neuroimage.2018.07.039

22. F-TRACT Consortium. F-TRACT: a probabilistic atlas of anatomo-functional connectivity of the human brain. EBRAINS. doi:10.25493/5AM4-J3F

23. Avalos-Alais S, Jedynak M, Boyer A, et al. High-resolution electrophysiological mapping of effective connectivity of lateral prefrontal cortex. Brain. Published online September 9, 2025. doi:10.1093/brain/awaf317

24. Lemarechal JD, Jedynak M, Trebaul L, et al. A brain atlas of axonal and synaptic delays based on modelling of cortico-cortical evoked potentials. Brain. 2022;145(5):1653–1667. doi:10.1093/brain/awab362

25. Deman P, Bhattacharjee M, Tadel F, et al. IntrAnat Electrodes: A Free Database and Visualization Software for Intracranial Electroencephalographic Data Processed for Case and Group Studies. Front Neuroinform. 2018;12. doi:10.3389/fninf.2018.00040

26. Guevara P, Duclap D, Poupon C, et al. Automatic fiber bundle segmentation in massive tractography datasets using a multi-subject bundle atlas. Neuroimage. 2012;61(4):1083–1099. doi:10.1016/j.neuroimage.2012.02.071

27. Amunts K, Mohlberg H, Bludau S, Zilles K. Julich-Brain: A 3D probabilistic atlas of the human brain’s cytoarchitecture. Science (80-). 2020;369(6506):988–992. doi:10.1126/science.abb4588

28. Hagmann P, Cammoun L, Gigandet X, et al. Mapping the Structural Core of Human Cerebral Cortex. Friston KJ, ed. PLoS Biol. 2008;6(7):e159. doi:10.1371/journal.pbio.0060159

29. Richardson JD, Fridriksson J. The Insular Cortex. In: Neurobiology of Language. Elsevier; 2016:115–127. doi:10.1016/B978-0-12-407794-2.00010-9

30. Cammoun L, Gigandet X, Meskaldji D, et al. Mapping the human connectome at multiple scales with diffusion spectrum MRI. J Neurosci Methods. 2012;203(2):386–397. doi:10.1016/j.jneumeth.2011.09.031

31. Tyler J. Torrico SA. Neuroanatomy, Limbic System. StatPearls Publishing; 2020. https://pubmed.ncbi.nlm.nih.gov/30860726/

32. Dancey CP, Reidy J. Statistics without Maths for Psychology. Pearson education; 2007.

33. Cohen J. Statistical Power Analysis for the Behavioral Sciences. Routledge; 2013. doi:10.4324/9780203771587

34. Roehri N, Pizzo F, Bartolomei F, Wendling F, Bénar C-G. What are the assets and weaknesses of HFO detectors? A benchmark framework based on realistic simulations. Avoli M, ed. PLoS One. 2017;12(4):e0174702. doi:10.1371/journal.pone.0174702

35. Evrard HC. The Organization of the Primate Insular Cortex. Front Neuroanat. 2019;13(May):1–21. doi:10.3389/fnana.2019.00043

36. Bastuji H, Frot M, Perchet C, Hagiwara K, Garcia-Larrea L. Convergence of sensory and limbic noxious input into the anterior insula and the emergence of pain from nociception. Sci Rep. 2018;8(1):13360. doi:10.1038/s41598-018-31781-z

37. Allen M, Fardo F, Dietz MJ, et al. Anterior insula coordinates hierarchical processing of tactile mismatch responses. Neuroimage. 2016;127:34–43. doi:10.1016/j.neuroimage.2015.11.030

38. Seth AK. Interoceptive inference, emotion, and the embodied self. Trends Cogn Sci. 2013;17(11):565–573. doi:10.1016/j.tics.2013.09.007

39. van den Heuvel MP, Kahn RS, Goñi J, Sporns O. High-cost, high-capacity backbone for global brain communication. Proc Natl Acad Sci. 2012;109(28):11372–11377. doi:10.1073/pnas.1203593109

40. Menon V, Uddin LQ. Saliency, switching, attention and control: a network model of insula function. Brain Struct Funct. 2010;214(5-6):655–667. doi:10.1007/s00429-010-0262-0

41. Sridharan D, Levitin DJ, Menon V. A critical role for the right fronto-insular cortex in switching between central-executive and default-mode networks. Proc Natl Acad Sci. 2008;105(34):12569–12574. doi:10.1073/pnas.0800005105

42. Huang W, Lyu D, Stieger JR, et al. Direct interactions between the human insula and hippocampus during memory encoding. Nat Neurosci. 2025;28(8):1763–1771. doi:10.1038/s41593-025-02005-1

43. Cerliani L, Thomas RM, Jbabdi S, et al. Probabilistic tractography recovers a rostrocaudal trajectory of connectivity variability in the human insular cortex. Hum Brain Mapp. 2012;33(9):2005–2034. doi:10.1002/hbm.21338

44. Deen B, Pitskel NB, Pelphrey KA. Three Systems of Insular Functional Connectivity Identified with Cluster Analysis. Cereb Cortex. 2011;21(7):1498–1506. doi:10.1093/cercor/bhq186

45. Biduła SP, Króliczak G. Structural asymmetry of the insula is linked to the lateralization of gesture and language. Eur J Neurosci. 2015;41(11):1438–1447. doi:10.1111/ejn.12888

46. Leonard MK, Cai R, Babiak MC, Ren A, Chang EF. The peri-Sylvian cortical network underlying single word repetition revealed by electrocortical stimulation and direct neural recordings. Brain Lang. 2019;193:58–72. doi:10.1016/j.bandl.2016.06.001

47. Shum M, Shiller DM, Baum SR, Gracco VL. Sensorimotor integration for speech motor learning involves the inferior parietal cortex. Eur J Neurosci. 2011;34(11):1817–1822. doi:10.1111/j.1460-9568.2011.07889.x

48. Raichle ME. The Brain’s Default Mode Network. Annu Rev Neurosci. 2015;38(1):433–447. doi:10.1146/annurev-neuro-071013-014030

49. KRINGELBACH M. The functional neuroanatomy of the human orbitofrontal cortex: evidence from neuroimaging and neuropsychology. Prog Neurobiol. 2004;72(5):341–372. doi:10.1016/j.pneurobio.2004.03.006

50. Dalenberg JR, Hoogeveen HR, Renken RJ, Langers DRM, ter Horst GJ. Functional specialization of the male insula during taste perception. Neuroimage. 2015;119:210–220. doi:10.1016/j.neuroimage.2015.06.062

51. Stephani C, Fernandez-Baca Vaca G, Maciunas R, Koubeissi M, Lüders HO. Functional neuroanatomy of the insular lobe. Brain Struct Funct. 2011;216(2):137–149. doi:10.1007/s00429-010-0296-3

52. Enatsu R, Gonzalez-Martinez J, Bulacio J, et al. Connectivity of the frontal and anterior insular network: A cortico-cortical evoked potential study. J Neurosurg. 2016;125(1):90–101. doi:10.3171/2015.6.JNS15622

53. Malinen S, Vartiainen N, Hlushchuk Y, et al. Aberrant temporal and spatial brain activity during rest in patients with chronic pain. Proc Natl Acad Sci. 2010;107(14):6493–6497. doi:10.1073/pnas.1001504107

54. Kucyi A, Moayedi M, Weissman-Fogel I, Hodaie M, Davis KD. Hemispheric asymmetry in white matter connectivity of the temporoparietal junction with the insula and prefrontal cortex. PLoS One. 2012;7(4). doi:10.1371/journal.pone.0035589

55. Chee MWL, Soon CS, Lee HL, Pallier C. Left insula activation: A marker for language attainment in bilinguals. Proc Natl Acad Sci. 2004;101(42):15265–15270. doi:10.1073/pnas.0403703101

56. Guenther FH. Cortical interactions underlying the production of speech sounds. J Commun Disord. 2006;39(5):350–365. doi:10.1016/j.jcomdis.2006.06.013

57. Oh A, Duerden EG, Pang EW. The role of the insula in speech and language processing. Brain Lang. 2014;135:96–103. doi:10.1016/j.bandl.2014.06.003

58. Fedorenko E, Ivanova AA, Regev TI. The language network as a natural kind within the broader landscape of the human brain. Nat Rev Neurosci. 2024;25(5):289–312. doi:10.1038/s41583-024-00802-4

59. Corbetta M, Patel G, Shulman GL. The Reorienting System of the Human Brain: From Environment to Theory of Mind. Neuron. 2008;58(3):306–324. doi:10.1016/j.neuron.2008.04.017

60. Wang Y, Yang X, Xiao L, et al. Altered anterior insula-superior frontal gyrus functional connectivity is correlated with cognitive impairment following total sleep deprivation. Biochem Biophys Res Commun. 2022;624:47–52. doi:10.1016/j.bbrc.2022.07.078

61. Kurth F, Eickhoff SB, Schleicher A, Hoemke L, Zilles K, Amunts K. Cytoarchitecture and probabilistic maps of the human posterior insular cortex. Cereb Cortex. 2010;20(6):1448–1461. doi:10.1093/cercor/bhp208

62. Cecchi R, Collomb-Clerc A, Rachidi I, et al. Direct stimulation of anterior insula and ventromedial prefrontal cortex disrupts economic choices. Nat Commun . 2024;15(1):1–11. doi:10.1038/s41467-024-51822-8

63. Yamao Y, Matsumoto R, Kunieda T, et al. Intraoperative dorsal language network mapping by using single pulse electrical stimulation. Hum Brain Mapp. 2014;35(9):4345–4361. doi:10.1002/hbm.22479

64. Keller CJ, Bickel S, Entz L, et al. Intrinsic functional architecture predicts electrically evoked responses in the human brain. Proc Natl Acad Sci. 2011;108(25):10308–10313. doi:10.1073/pnas.1019750108

65. Turpin C, Rossel O, Schlosser-Perrin F, et al. Shapes of direct cortical responses vs. short-range axono-cortical evoked potentials: The effects of direct electrical stimulation applied to the human brain. Clin Neurophysiol. 2025;169:91–99. doi:10.1016/j.clinph.2024.10.016

66. Valentin A. Responses to single pulse electrical stimulation identify epileptogenesis in the human brain in vivo. Brain. 2002;125(8):1709–1718. doi:10.1093/brain/awf187

67. Donos C, Mîndruţă I, Malîia MD, Raşină A, Ciurea J, Barborica A. Co-occurrence of high-frequency oscillations and delayed responses evoked by intracranial electrical stimulation in stereo-EEG studies. Clin Neurophysiol. 2017;128(6):1043–1052. doi:10.1016/j.clinph.2016.11.028

68. Boido D, Kapetis D, Gnatkovsky V, et al. Stimulus evoked potentials contribute to map the epileptogenic zone during stereo EEG presurgical monitoring. Hum Brain Mapp. 2014;35(9):4267–4281. doi:10.1002/hbm.22516

69. Lacruz ME, García Seoane JJ, Valentin A, Selway R, Alarcón G. Frontal and temporal functional connections of the living human brain. Eur J Neurosci. 2007;26(5):1357–1370. doi:10.1111/j.1460-9568.2007.05730.x

70. Ryvlin P, Minotti L, Demarquay G, et al. Nocturnal Hypermotor Seizures, Suggesting Frontal Lobe Epilepsy, Can Originate in the Insula. Epilepsia. 2006;47(4):755–765. doi:10.1111/j.1528-1167.2006.00510.x

71. Terasawa Y, Shibata M, Moriguchi Y, Umeda S. Anterior insular cortex mediates bodily sensibility and social anxiety. Soc Cogn Affect Neurosci. 2013;8(3):259–266. doi:10.1093/scan/nss108

72. Wylie KP, Tregellas JR. The role of the insula in schizophrenia. Schizophr Res. 2010;123(2-3):93–104. doi:10.1016/j.schres.2010.08.027

73. Blair RJR. The neurobiology of psychopathic traits in youths. Nat Rev Neurosci. 2013;14(11):786–799. doi:10.1038/nrn3577

74. Kaye WH, Fudge JL, Paulus M. New insights into symptoms and neurocircuit function of anorexia nervosa. Nat Rev Neurosci. 2009;10(8):573–584. doi:10.1038/nrn2682

75. Löffler LAK, Radke S, Morawetz C, Derntl B. Emotional dysfunctions in neurodegenerative diseases. J Comp Neurol. 2016;524(8):1727–1743. doi:10.1002/cne.23816

76. Bonthius DJ, Solodkin A, Van Hoesen GW. Pathology of the Insular Cortex in Alzheimer Disease Depends on Cortical Architecture. J Neuropathol Exp Neurol. 2005;64(10):910–922. doi:10.1097/01.jnen.0000182983.87106.d1

77. Uddin LQ, Supekar K, Lynch CJ, et al. Brain State Differentiation and Behavioral Inflexibility in Autism. Cereb Cortex. 2015;25(12):4740–4747. doi:10.1093/cercor/bhu161

78. Kühn S, Gallinat J. Common biology of craving across legal and illegal drugs - a quantitative meta-analysis of cue-reactivity brain response. Eur J Neurosci. 2011;33(7):1318–1326. doi:10.1111/j.1460-9568.2010.07590.x

