## Supplementary Material for "Effective connectivity of the insula as measured by cortico-cortical evoked potentials"

### Materials and methods

#### 1. SEEG acquisition

Details on the anatomical and functional preprocessing steps to compute the CCEPs are available in two previous studies[1,2]. In summary, electrode and contact positions extracted from the post-implantation CT scan were co-registered to the pre-implantation 3D T1 MRI of the participant and their tissue classification was determined according to grey/white matter segmentation of the MRI. Then, spatial normalization projected the contacts of each patient in the MNI referential. IntranatElectrodes software allowed, then, to compute region labels with respect to a series of neuroanatomical atlases for each contact.

Regarding SEEG data, bad channels were pre-identified with a machine learning approach and then visual inspected by SEEG experts for final classification. Data were re-referenced with a bipolar montage of adjacent contacts to retain only focal activities. Stimulation runs (a row of single electrical pulses) were automatically detected based on stimulation artifacts. These transient artifacts were corrected using an interpolation surrounding the artefact's peak. Then, the data were band-pass filtered between 1 and 45 Hz. Epochs were defined on a [-200 800] ms interval around the stimulation pulse and averaged after removing outlier pulses (i.e., pulses related to responses 3 times higher than the median response over the stimulation run). Finally, CCEPs were z-scored with respect to [-200 -10] ms baseline preceding each stimulation pulse. The software suite used to perform these preprocessing steps is open-source available from the F-TRACT project at f-tract.eu/software.

### 2. Maximum peak delay

Figure 1 shows the Spearman correlation coefficient, *r*, between probability matrices generated from different maximum peak delays. Probability matrices were generated considering Montreal1 stimulation and Lausanne-scale2 recording schemes. Very strong correlations are considered for $r > 0.9$ [4]. By analyzing Figure 2, we see that connectivity matrices derived from different time windows were strongly correlated ($r > 0.93$), indicating strong consistency across window choices.

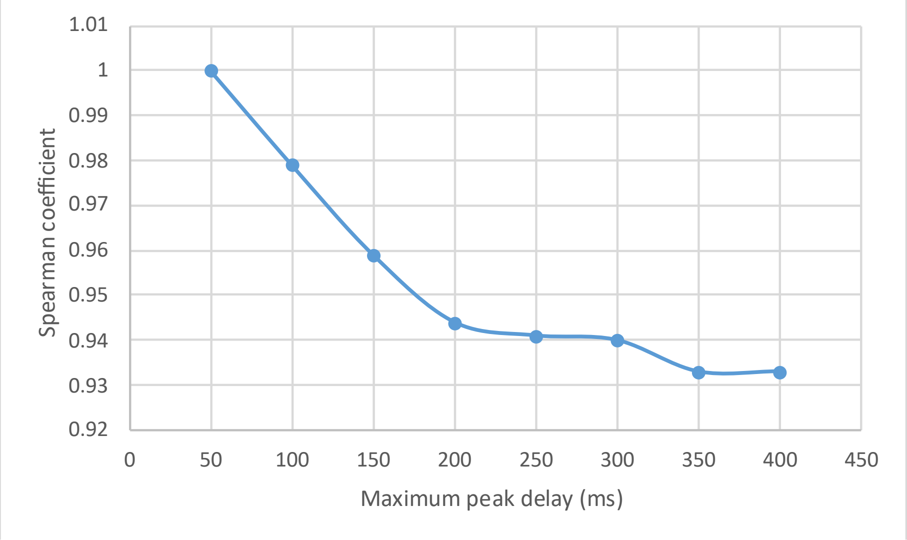


**Figure 1 –** Spearman correlation coefficient between probability matrices generated from different maximum peak delays (50-400ms) against a probability matrix generated from a maximum peak delay of 50ms.

#### 3. Minimum responses to compute connectivity outcomes

Brain coverage is calculated as the percentage of recording brain regions that accomplish the minimum required responses in the total number of brain regions according to an atlas scheme. Only the same hemisphere as the stimulation region is accounted for intra-hemisphere coverage. The confidence interval was calculated according to Agresti-Coull formula[3] considering a significance level of 0.05.

**Table 1** – Intra-hemisphere and inter-hemisphere coverage (%) and confidence interval of probability of effective connectivity for Montreal1 stimulation and Lausanne-scale2 recording schemes using the requirement of 10, 50, and 90 minimum responses

| Minimum responses | % Intra-hemisphere coverage | % Inter-hemisphere coverage | Average confidence interval (min-max) |
|---|---|---|---|
| 10 | 93 | 86 | 0.02 (0.005-0.23) |
| 50 | 89 | 86 | 0.02 (0.005-0.10) |
| 90 | 89 | 81 | 0.02 (0.005-0.08) |

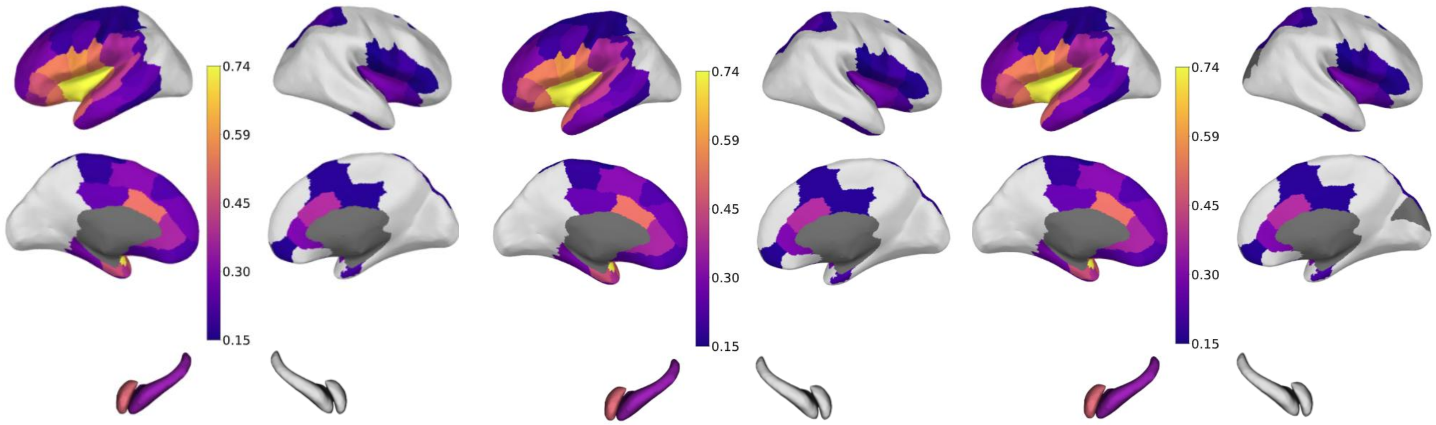


(A) (B) (C)

**Figure 2** – Matrices of probability of effective connectivity for Montreal1 stimulation and Lausanne-scale2 recording schemes using the requirement of **(A)** 10, **(B)** 50, and **(C)** 90 minimum responses. Grey cells correspond to regions in which the total number of responses are less than the value required.

**Table 2** – Intra-hemisphere coverage (%) and probability's confidence interval using the requirement of 50 minimum responses to compute connectivity outcomes for each pair stimulated and recorded atlas scheme

| Stimulation & recording atlas schemes | % coverage | Average confidence interval (min-max) |
|---|---|---|
| Montreal1 & Lausanne-scale2 | 89 | 0.02 (0.005-0.10) |
| Montreal4 & Lausanne-scale2 | 85 | 0.03 (0.007-0.13) |
| Montreal19 & Lausanne-scale2 | 76 | 0.05 (0.012-0.13) |
| Lausanne-scale2 & Montreal1 | 90 | 0.02 (0.005-0.12) |

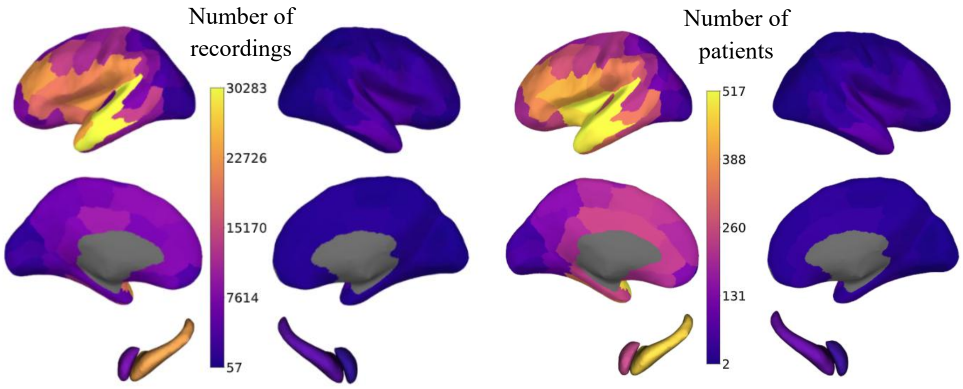


**(A)** **(B)**

**Figure 3** – Number of **(A)** recordings and **(B)** patients considering the requirement of 50 minimum responses from at least 2 patients.

### 4. Surrogate threshold

Surrogate probabilities were generated multiple times from fake stimulation events randomly distributed during pre-stimulation period. To define the surrogate threshold, as we have done in[2], the same algorithm was used to identify the CCEPs on a subset of our data but with events randomly distributed in the time series recorded during the period before stimulation, called "surrogate events". In each run, the number of surrogate events matched the number of stimulation pulses. Baseline signals from before surrogate events [-200, -10] ms were used to correct and z-score the surrogate signal. For an one-side test, the surrogate threshold, for a given p-value, corresponds to the nth value of the increasing ranked surrogate distribution given by the formula: p-value = 1-(n/(N+1)) where N is the total number of surrogates[11].

### 5. Atlas schemes

#### 5.1. Insula

Figures 4A and 5A show the left insula from Montreal and Julich atlas schemes, respectively. Both atlas schemes are symmetric regarding hemispheres. Montreal's scheme comprises 19 insula parcels labeled with "R" or "L" for right and left hemisphere, respectively, followed by the number 1-19[5]. Julich's scheme includes 16 insula parcels per hemisphere labeled with "Id", "Ia", or "Ig" for left dysgranular, agranular, or granular regions, respectively, followed by a number 1-10, 1-3, or 1-3, respectively[6,7].

Figures 4B and 5B present insula with 4 parcel resolution considering Montreal and Julich schemes, respectively. Montreal4 was computed based on the classification defined in Table 3 of Ghaziri et al.[5] whereas Julich4 was derived from the cytoarchitectonic cluster analysis presented in Quabs et al.[6], resulting in the following insula subregions: anterior superior (blue), anterior inferior (grey), posterior superior (green), and posterior inferior (orange).

Mid-to-high resolution (Figures 4C and 5C) was developed based on the anatomical boundaries defined by the mid resolution scheme. Montreal7 was constructed according to the sulco-gyral anatomy described in [5,8,9]. In summary, the division between anterior and posterior regions was defined by the insular central sulcus. Within the anterior superior insula, three subregions were identified based on the anterior and precentral sulci: the short anterior insular gyrus, short middle insular gyrus, and short posterior insular gyrus. The posterior superior insula was subdivided into two regions separated by the postcentral sulcus: the long anterior insular gyrus and the long posterior insular gyrus. Julich10 was derived to balance increased spatial resolution with sufficient coverage according to F-TRACT data density, preserving consistency with the underlying cytoarchitectonic division from Quabs et al.[6].

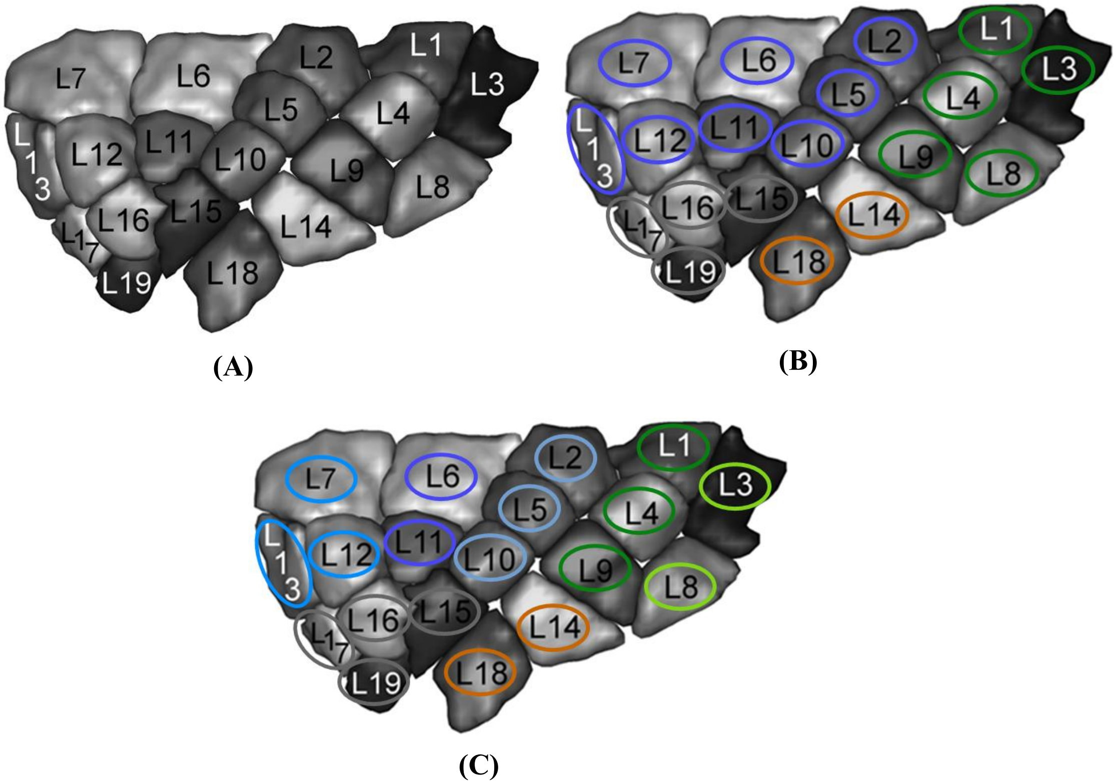


**Figure 4** – Levels of Montreal atlas resolution: **(A)** high resolution with 19 insula parcels (Montreal19); **(B)** mid resolution with 4 insula parcels (Montreal4); **(C)** mid-to-high resolution with 7 insula parcels (Montreal7). Adapted from[5].

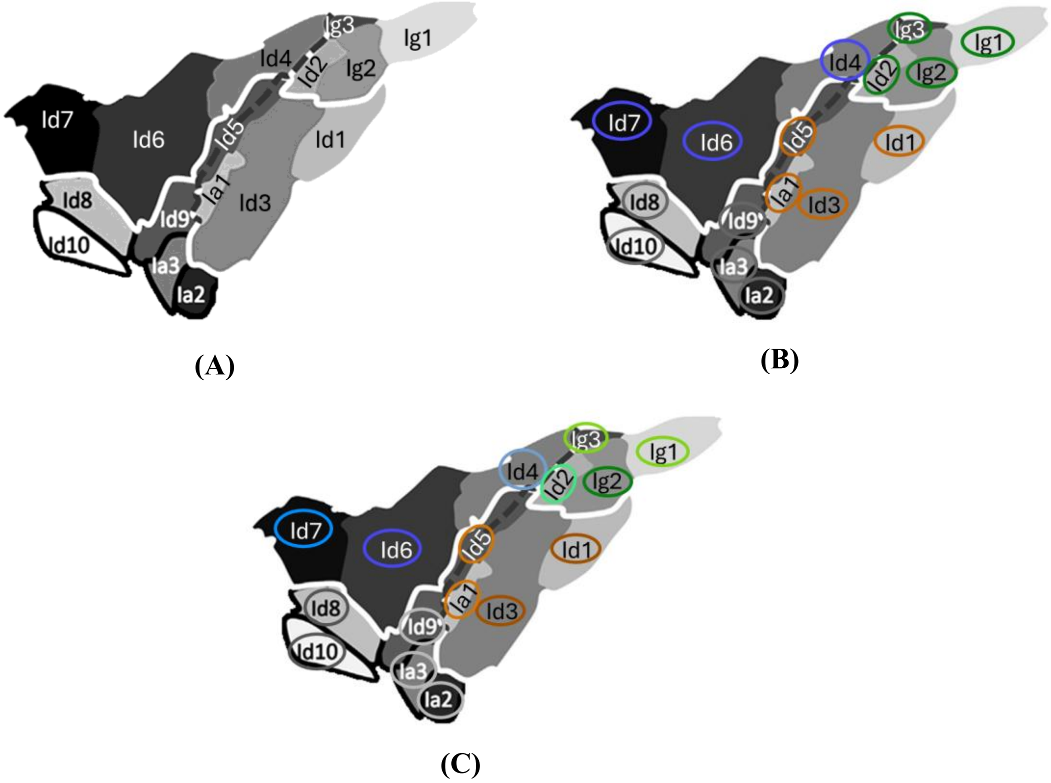


**Figure 5** – Levels of Julich atlas resolution: **(A)** high resolution with 16 insula parcels (Julich16); **(B)** mid resolution with 4 insula parcels (Julich4); **(C)** mid-to-high resolution with 10 insula parcels (Julich10). Adapted from [6,7].

### 5.2.Whole brain

Table 3 describes the labels of parcels from Lausanne2018-scale2 atlas scheme, and their corresponding lobe and system (occipital, temporal, parietal, central, frontal, or limbic) according to Desikan et al.[10]. Figure 6 demonstrates the position of each parcel on a brain map.

**Table 1** – Labels of Lausanne2018-scale2's parcels and corresponding brain lobe and system. An acronym was also defined for each parcel

| Left Hemisphere Label | Right Hemisphere Label | Acronym | System | Lobe |
|---|---|---|---|---|
| lh.cuneus_1 | rh.cuneus_1 | Cu | Occipital | Occipital |
| lh.lateraloccipital_1 | rh.lateraloccipital_1 | LO1 | Occipital | Occipital |
| lh.lateraloccipital_2 | rh.lateraloccipital_2 | LO2 | Occipital | Occipital |
| lh.lateraloccipital_3 | rh.lateraloccipital_3 | LO3 | Occipital | Occipital |
| lh.lingual_1 | rh.lingual_1 | L1 | Occipital | Occipital |
| lh.lingual_2 | rh.lingual_2 | L2 | Occipital | Occipital |
| lh.pericalcarine_1 | rh.pericalcarine_1 | P | Occipital | Occipital |
| lh.bankssts_1 | rh.bankssts_1 | B | Temporal | Temporal |
| lh.entorhinal_1 | rh.entorhinal_1 | E | Limbic | Temporal |
| lh.fusiform_1 | rh.fusiform_1 | F1 | Temporal | Temporal |
| lh.fusiform_2 | rh.fusiform_2 | F2 | Temporal | Temporal |
| lh.inferiortemporal_1 | rh.inferiortemporal_1 | IT1 | Temporal | Temporal |
| lh.inferiortemporal_2 | rh.inferiortemporal_2 | IT2 | Temporal | Temporal |
| lh.middletemporal_1 | rh.middletemporal_1 | MT1 | Temporal | Temporal |
| lh.middletemporal_2 | rh.middletemporal_2 | MT2 | Temporal | Temporal |
| lh.parahippocampal_1 | rh.parahippocampal_1 | PH | Limbic | Temporal |
| lh.superiortemporal_1 | rh.superiortemporal_1 | ST1 | Temporal | Temporal |
| lh.superiortemporal_2 | rh.superiortemporal_2 | ST2 | Temporal | Temporal |
| lh.temporalpole_1 | rh.temporalpole_1 | TP | Limbic | Temporal |
| lh.transversetemporal_1 | rh.transversetemporal_1 | TT | Temporal | Temporal |
| lh.inferiorparietal_1 | rh.inferiorparietal_1 | IP1 | Parietal | Parietal |
| lh.inferiorparietal_2 | rh.inferiorparietal_2 | IP2 | Parietal | Parietal |
| lh.inferiorparietal_3 | rh.inferiorparietal_3 | IP3 | Parietal | Parietal |
| lh.postcentral_1 | rh.postcentral_1 | PoC1 | Central | Parietal |
| lh.postcentral_2 | rh.postcentral_2 | PoC2 | Central | Parietal |
| lh.precuneus_1 | rh.precuneus_1 | PCu1 | Parietal | Parietal |
| lh.precuneus_2 | rh.precuneus_2 | PCu2 | Parietal | Parietal |
| lh.superiorparietal_1 | rh.superiorparietal_1 | SP1 | Parietal | Parietal |
| lh.superiorparietal_2 | rh.superiorparietal_2 | SP2 | Parietal | Parietal |
| lh.superiorparietal_3 | rh.superiorparietal_3 | SP3 | Parietal | Parietal |
| lh.supramarginal_1 | rh.supramarginal_1 | SM1 | Parietal | Parietal |
| lh.supramarginal_2 | rh.supramarginal_2 | SM2 | Parietal | Parietal |
| lh.caudalanteriorcingulate_1 | rh.caudalanteriorcingulate_1 | CAC | Limbic | Frontal |
| lh.isthmuscingulate_1 | rh.isthmuscingulate_1 | IC | Parietal | Parietal |
| lh.posteriorcingulate_1 | rh.posteriorcingulate_1 | PC | Parietal | Parietal |

| | | | | |
|---|---|---|---|---|
| lh.rostralanteriorcingulate_1 | rh.rostralanteriorcingulate_1 | RAC | Limbic | Frontal |
| lh.paracentral_1 | rh.paracentral_1 | PaC | Central | Parietal |
| lh.caudalmiddlefrontal_1 | rh.caudalmiddlefrontal_1 | CMF | Frontal | Frontal |
| lh.frontalpole_1 | rh.frontalpole_1 | FP | Frontal | Frontal |
| lh.parsopercularis_1 | rh.parsopercularis_1 | POp | Frontal | Frontal |
| lh.parstriangularis_1 | rh.parstriangularis_1 | PT | Frontal | Frontal |
| lh.precentral_1 | rh.precentral_1 | PrC1 | Central | Frontal |
| lh.precentral_2 | rh.precentral_2 | PrC2 | Central | Frontal |
| lh.precentral_3 | rh.precentral_3 | PrC3 | Central | Frontal |
| lh.rostralmiddlefrontal_1 | rh.rostralmiddlefrontal_1 | RMF1 | Frontal | Frontal |
| lh.rostralmiddlefrontal_2 | rh.rostralmiddlefrontal_2 | RMF2 | Frontal | Frontal |
| lh.superiorfrontal_1 | rh.superiorfrontal_1 | SF1 | Frontal | Frontal |
| lh.superiorfrontal_2 | rh.superiorfrontal_2 | SF2 | Frontal | Frontal |
| lh.superiorfrontal_3 | rh.superiorfrontal_3 | SF3 | Frontal | Frontal |
| lh.superiorfrontal_4 | rh.superiorfrontal_4 | SF4 | Frontal | Frontal |
| lh.lateralorbitofrontal_1 | rh.lateralorbitofrontal_1 | LOF1 | Limbic | Frontal |
| lh.lateralorbitofrontal_2 | rh.lateralorbitofrontal_2 | LOF2 | Limbic | Frontal |
| lh.medialorbitofrontal_1 | rh.medialorbitofrontal_1 | MOF1 | Limbic | Frontal |
| lh.medialorbitofrontal_2 | rh.medialorbitofrontal_2 | MOF2 | Limbic | Frontal |
| lh.parsorbitalis_1 | rh.parsorbitalis_1 | POr | Frontal | Frontal |
| lh.insula_1 | rh.insula_1 | I1 | - | - |
| lh.insula_2 | rh.insula_2 | I2 | - | - |
| lh.Hippocampus | rh.Hippocampus | H | Limbic | Temporal |
| lh.Amygdala | rh.Amygdala | A | Limbic | Temporal |

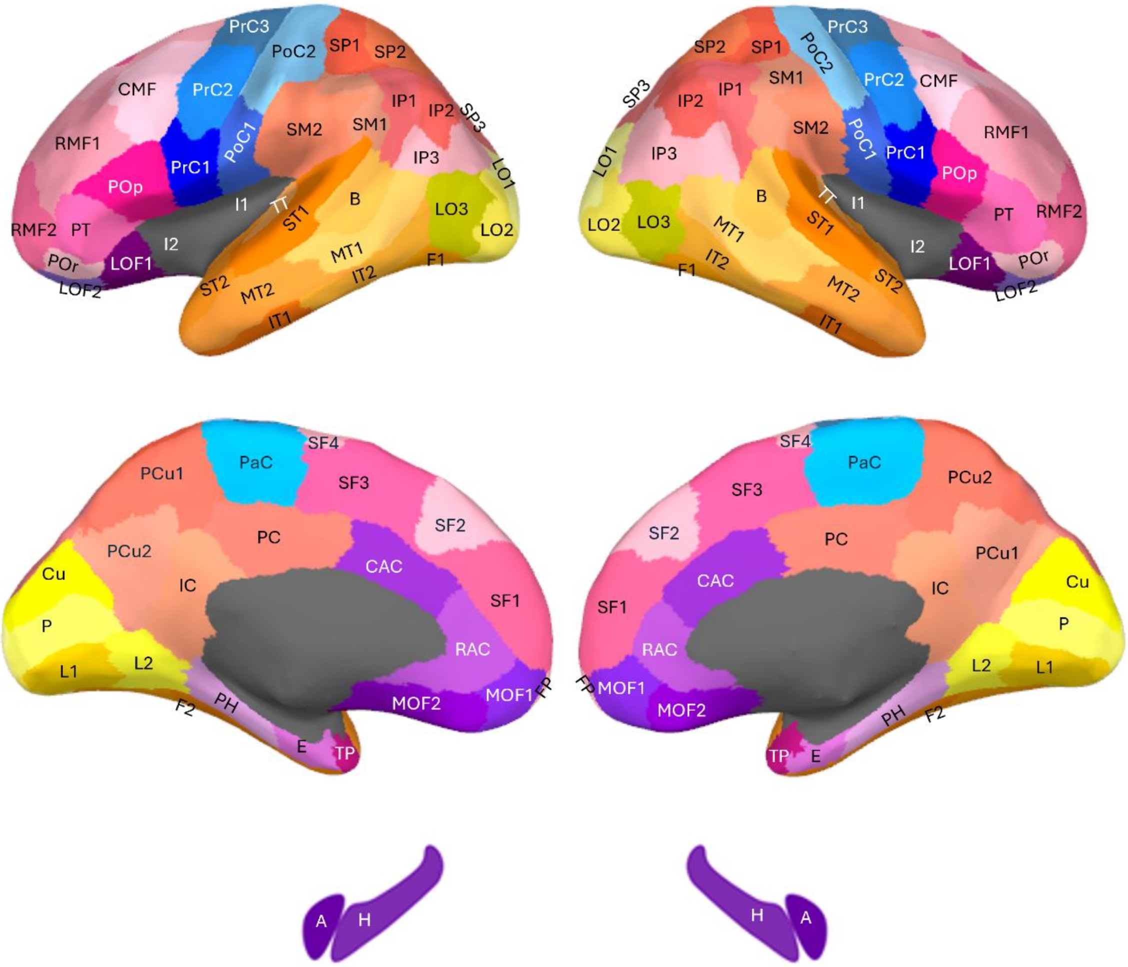


**Figure 6 –** Position of Lausanne2018-scale2 parcels on a brain map. Each parcel is identified by acronyms from Table 3. Parcels from central, parietal, temporal, occipital, frontal, and limbic systems are colored in blue, pink-orange, orange, yellow, pink, and purple, respectively.

**Results**

### 6. Results from Montreal atlas scheme

Response amplitude corresponded to the magnitude of the first peak above the z-score threshold. Figure 7 presents the median amplitude of connectivity between insula and the whole brain. The values of amplitude ranged between 7 and 15 z-score and showed a strongly correlated pattern with probability (r=0.81 and p-value=2.10e-24).

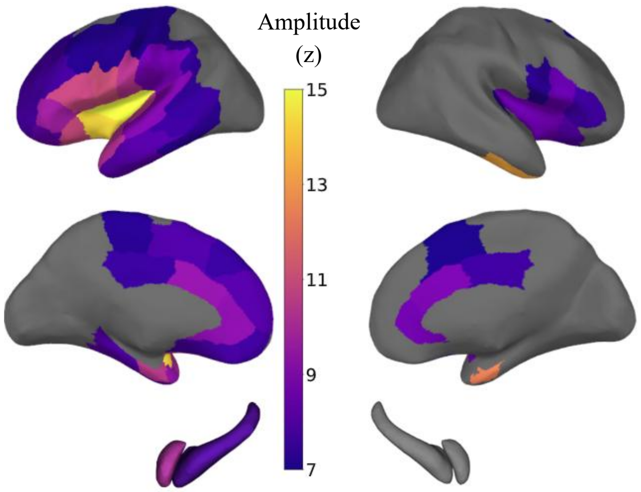


**Figure 7** – Median amplitude (z) for connectivity between Montreal1 stimulations and Lausanne-scale2 recordings. No color appears if the requirement for minimum number of responses is not met.

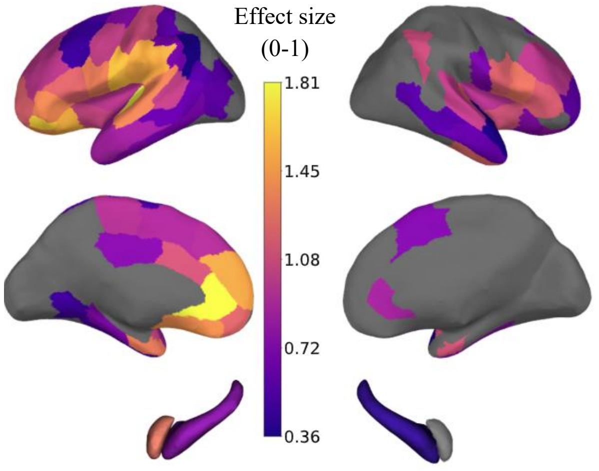


**Figure 8 –** Effect size [0,1] summarizing differences in whole-brain connectivity profiles across the 19 insular subregions.

### 7. Results from Jülich atlas scheme

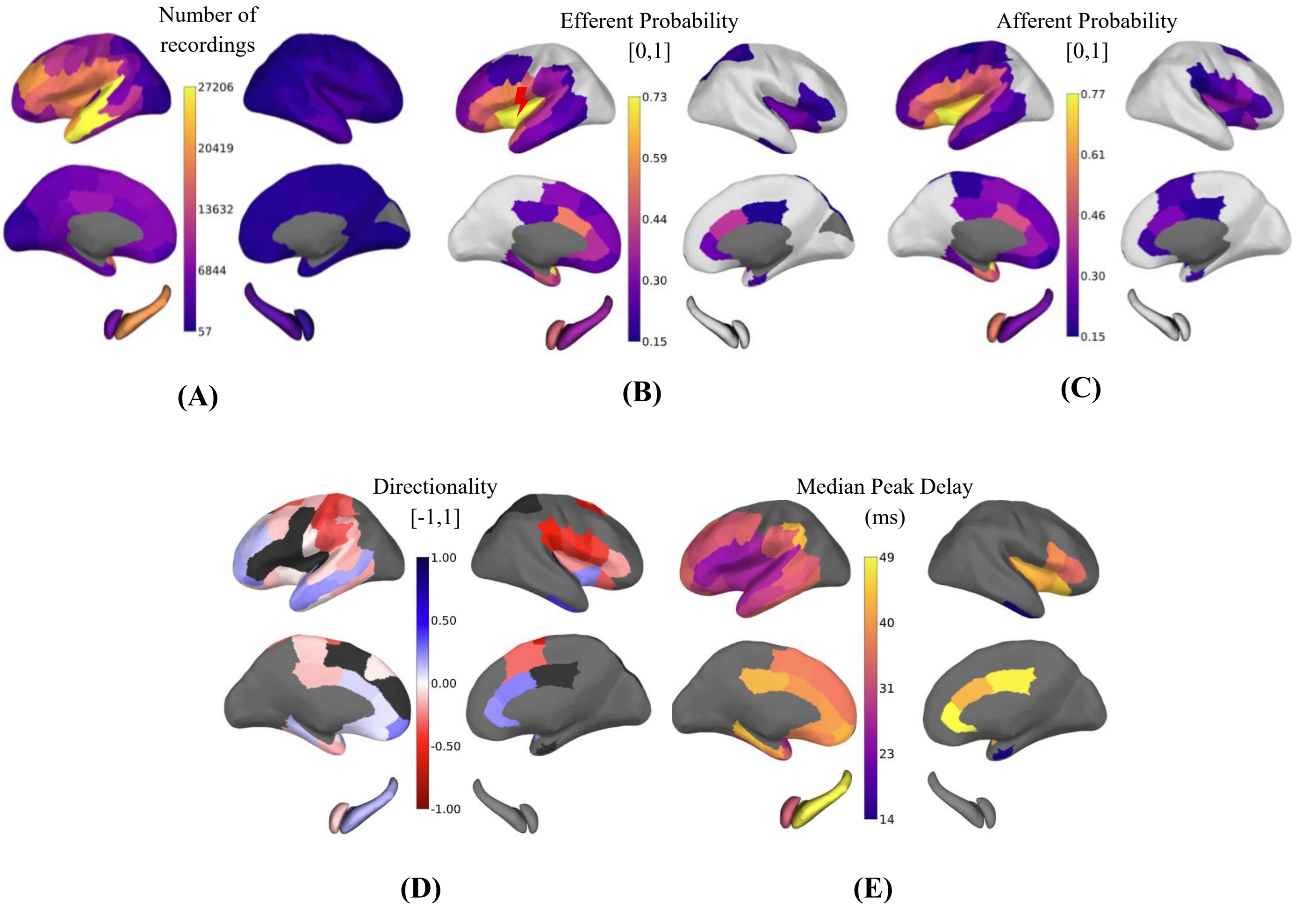


**Figure 9 – Connectivity of whole insula (Julich1). (A)** Number of recordings; **(B)** Efferent connectivity probability; **(C)** Afferent connectivity probability; **(D)** Directionality; **(E)** Median peak delay (ms). Light grey areas in (B-C) represent probabilities below surrogate threshold. No color appears if the requirement for minimum number of responses is not met. Dark grey color in (D) represents no statistically significant differences between efferent and afferent probabilities (p-value > 0.05).

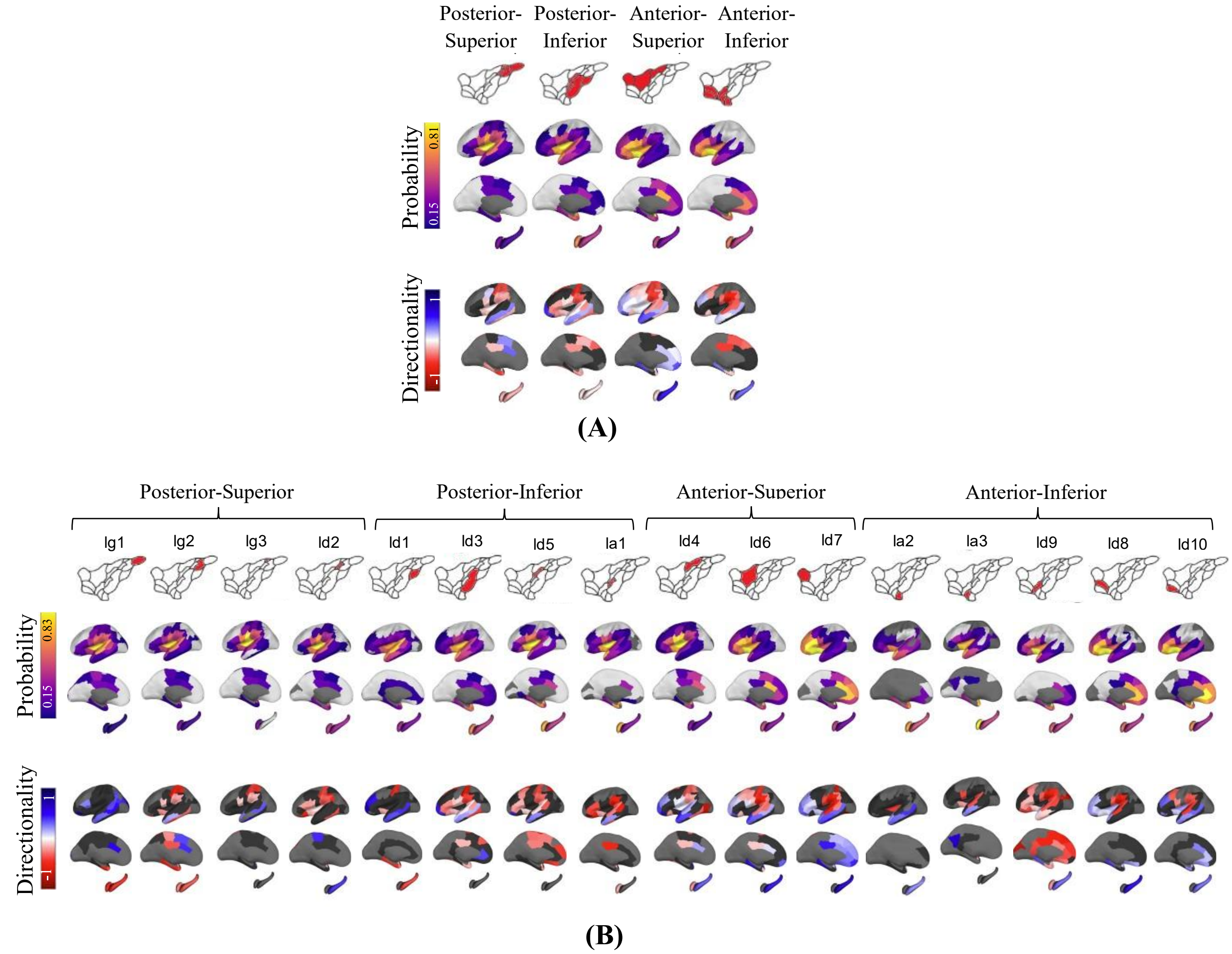


**Figure 10 - Insula connectivity at mid (A) and high (B) resolution.** Efferent probabilistic connectivity and directionality of effective connectivity for **(A)** Julich4 stimulations and **(B)** Julich16 stimulations. Light grey color represents probabilities below surrogate threshold. No color appears when the requirement for minimum number of responses is not met. Black color represents no statistically significant differences between efferent and afferent probabilities (p-value > 0.05).

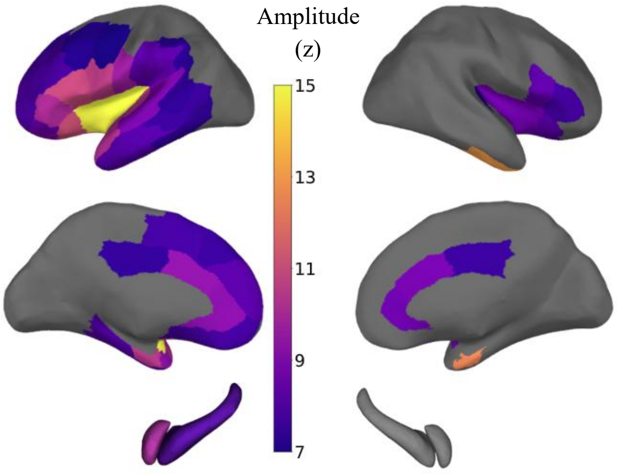


**Figure 1** – Median amplitude (z) for connectivity between Julich1 stimulations and Lausanne-scale2 recordings. No color appears if the requirement for minimum number of responses is not met.

By correlating probability matrices from Montreal and Jülich insula stimulation, we found a correlation coefficient of 0.99 (p-value=1.1e-98). Considering insula mid-resolution, we have an average of r=0.94±0.03 (Post-Sup: r=0.97 and p-value=5.0e-64; Post-Inf: r=0.89 and p-value=3.2e-33; Ant-Sup: r=0.98 and p-value=6.2e-74; Ant-Inf: r=0.92 and p-value=2.4e-37). For amplitude and peak delay, we found r=0.97 (p-value=5.8e-60) and r=0.96 (p-value=6.7e-56), respectively. These results suggest that the pattern of insular connectivity with the whole brain is consistent between both Montreal and Jülich atlas schemes.

### 8. Control results

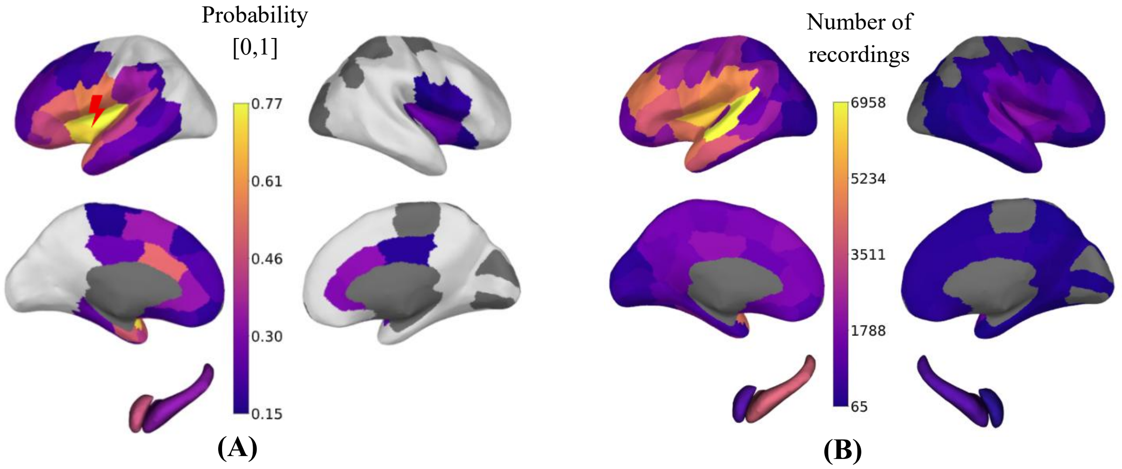


**Figure 2 – (A)** Probability of effective connectivity [0,1] and **(B)** number of recordings for Montreal1 stimulations and Lausanne-scale2 recordings using only CCEPs with zero spike count. White color represents probabilities below surrogate threshold. No color appears if the requirement for minimum number of responses is not met.

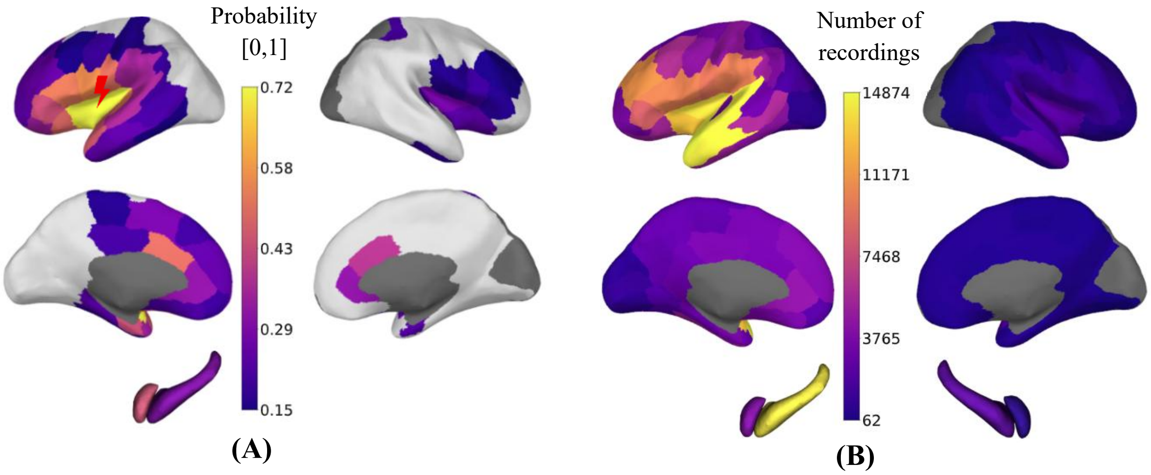


**Figure 13 – (A)** Probability of effective connectivity [0,1] and **(B)** number of recordings for Montreal1 stimulations and Lausanne-scale2 recordings using only contacts placed in grey matter. White color represents probabilities below surrogate threshold. No color appears if the requirement for minimum number of responses is not met.

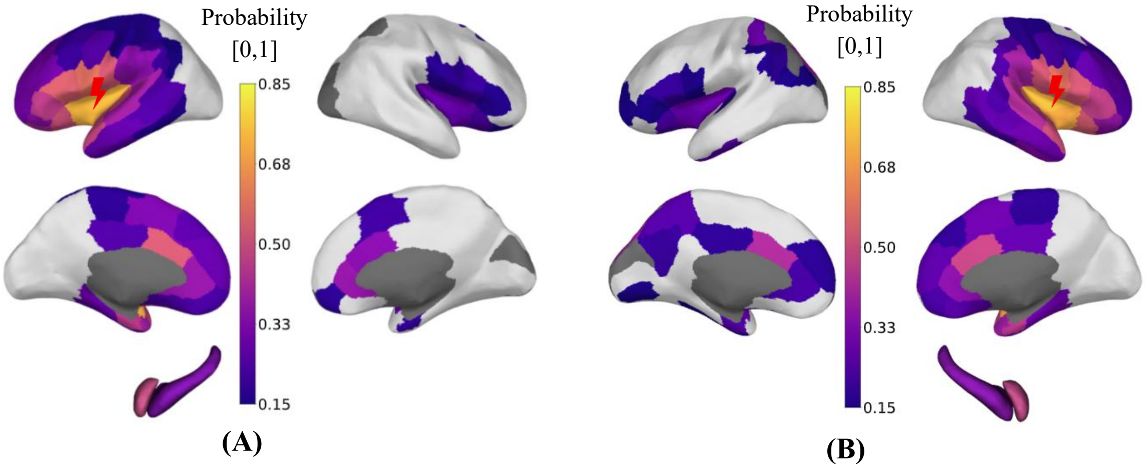


**Figure 14** – Probability of effective connectivity [0,1] for Montreal1 **(A)** left and **(B)** right hemisphere stimulations and Lausanne-scale2 recordings. White color represents probabilities below surrogate threshold. No color appears if the requirement for minimum number of responses is not met.

**Supplementary references**


1. David O, Job AS, De Palma L, Hoffmann D, Minotti L, Kahane P. Probabilistic functional tractography of the human cortex. *Neuroimage*. 2013;80:307-317. doi:10.1016/j.neuroimage.2013.05.075

2. Trebaul L, Deman P, Tuyisenge V, et al. Probabilistic functional tractography of the human cortex revisited. *Neuroimage*. 2018;181(June):414-429. doi:10.1016/j.neuroimage.2018.07.039

3. Agresti A, Coull BA. Approximate is Better than "Exact" for Interval Estimation of Binomial Proportions. *Am Stat*. 1998;52(2):119-126. doi:10.1080/00031305.1998.10480550

4. Dancey CP, Reidy J. *Statistics without Maths for Psychology*. Pearson education; 2007.

5. Ghaziri J, Tucholka A, Girard G, et al. The Corticocortical Structural Connectivity of the Human Insula. *Cereb Cortex*. 2017;27(2):1216-1228. doi:10.1093/cercor/bhv308

6. Quabs J, Caspers S, Schöne C, et al. Cytoarchitecture, probability maps and segregation of the human insula. *Neuroimage*. 2022;260(January):119453. doi:10.1016/j.neuroimage.2022.119453

7. Amunts K, Mohlberg H, Bludau S, Zilles K. Julich-Brain: A 3D probabilistic atlas of the human brain's cytoarchitecture. *Science (80- )*. 2020;369(6506):988-992. doi:10.1126/science.abb4588

8. Türe U, Yaşargil DCH, Al-Mefty O, Yaşargil MG. Topographic anatomy of the insular region. *J Neurosurg*. 1999;90(4):720-733. doi:10.3171/jns.1999.90.4.0720

9. Richardson JD, Fridriksson J. The Insular Cortex. In: *Neurobiology of Language*. Elsevier; 2016:115-127. doi:10.1016/B978-0-12-407794-2.00010-9

10. Desikan RS, Ségonne F, Fischl B, et al. An automated labeling system for subdividing the human cerebral cortex on MRI scans into gyral based regions of interest. *Neuroimage*. 2006;31(3):968-980. doi:10.1016/j.neuroimage.2006.01.021

11. Schreiber T, Schmitz A. Surrogate time series. *Phys D Nonlinear Phenom*. 2000;142(3-4):346-382. doi:10.1016/S0167-2789(00)00043-9
